# How does adversity in childhood relate to pain in adulthood? Findings across four experimental tests of pain modulation

**DOI:** 10.1101/2025.09.16.25335844

**Authors:** Gillian J Bedwell, Luyanduthando Mqadi, Mark R Hutchinson, Romy Parker, Victoria J Madden

## Abstract

Childhood adversity is commonly linked to persistent pain in adulthood. Altered nociceptive modulation may play a role in this relationship. We investigated whether childhood adversity is associated with four measures of experimental pain, and whether these associations differ by sex. In this experimental observational study, we enrolled pain-free adults with varying severity of childhood adversity, scored on the Childhood Trauma Questionnaire-Short Form. Experimental pain outcomes included the surface area and magnitude of induced secondary hyperalgesia, temporal summation, and conditioned pain modulation. We used linear regression to investigate whether childhood adversity was associated with increased surface area or magnitude of secondary hyperalgesia, increased temporal summation, or reduced conditioned pain modulation, and whether these relationships differed by sex. Ninety-five participants (61 female; median age 23 [IQR: 9] years) completed the study. Childhood adversity was positively associated with the surface area of secondary hyperalgesia (β=0.32 [95%CI: 0.01; 0.62], *p* = 0.04); however, this relationship was not upheld when accounting for multiple testing (*q* = 0.17). Childhood adversity was unrelated to the magnitude of secondary hyperalgesia, temporal summation, or conditioned pain modulation. These data suggest a subtle relationship between childhood adversity and the spatial extent of experimentally induced secondary hyperalgesia.

## Introduction

Adults with a history of adverse childhood experiences may have an elevated risk of persistent pain [7, 14, 21, 54, 69], greater pain severity [10, 15, 22, 54], and a wider anatomical distribution of persistent pain [15, 53]. Altered regulation of nociception may underlie this relationship: pain-free individuals with a history of childhood adversity display larger areas of capsaicin-induced secondary allodynia [72], greater temporal summation, and slower decay of temporal summation than individuals with no or minimal childhood adversity [71]. These data suggest that childhood adversity is linked to alterations in central pain modulation, which may confer risk of persistent pain.

Common experimental tests of central pain modulation include temporal summation, induced secondary hyperalgesia, and conditioned pain modulation (CPM). Assessments of temporal summation and induced secondary hyperalgesia provide insight to central facilitation of nociception. Temporal summation is thought to be mediated by homotopic facilitation in the dorsal horn of the spinal cord [44]. In contrast, secondary hyperalgesia is thought to be mediated by heterotopic facilitation [35, 45] and may be a function of both facilitatory and inhibitory nociceptive processes. During induction of secondary hyperalgesia, functional magnetic resonance imaging shows reduced activity of supraspinal regions involved in descending inhibition of nociception, suggesting that induced secondary hyperalgesia is mediated by not only increased facilitation but also decreased inhibition of nociception [48]. Secondary hyperalgesia can be experimentally induced using high-frequency electrical stimulation [35, 47, 61] and assessments of the surface area and magnitude of the induced secondary hyperalgesia are thought to represent nociceptive synaptic hyperexcitability in the spinal dorsal horn [27, 33–35, 45]. Assessment of CPM provides insight to central inhibition of nociception [8, 9, 38]. Therefore, assessing temporal summation, induced secondary hyperalgesia, and CPM in the same cohort offers deeper insight into nociceptive processing than evaluating each alone.

Responses to experimental pain challenges may differ between sex. Compared to males, females show larger areas of thermally induced secondary hyperalgesia, faster nociceptive withdrawal reflexes, and reduced CPM [20, 24]. One study found that pain-free females with a history of childhood adversity display greater pain sensitivity than females without childhood adversity; however, no such relationship was found in males [18]. Taken together, these data suggest that the relationship between childhood adversity and experimental pain outcomes may differ between males and females.

In a cohort of pain-free adults with varying severity of childhood adversity, we investigated whether childhood adversity was positively associated with (1) the surface area and (2) the magnitude of induced secondary hyperalgesia and (3) temporal summation, and negatively associated with (4) CPM. We also aimed to investigate differences and similarities between males and females for each of these relationships.

## Methods

### Study overview

This secondary analysis used data from an experimental study in humans [3], which was approved by the University of Cape Town, Faculty of Health Sciences Human Research Ethics Committee (560/2021), registered at clinicaltrials.gov (NCT06127693), and locked online at Open Science Framework https://osf.io/392zh/. We followed the STROBE reporting guidelines [62] (Supplementary file: Section 1, Table S1). The results were presented to and discussed with a group of people with a lived experience of persistent pain to learn about their perspectives on the key message(s) of this work and to obtain advice on strategies to disseminate these messages to the general public.

Generally healthy, pain-free adult participants with a range of self-reported childhood adversity underwent assessments of temporal summation and CPM in the lumbar region and at the deltoid insertion, and induced secondary hyperalgesia at one forearm. The primary study from which these data were obtained assessed temporal summation and CPM at two anatomical sites, the lumbar region and the deltoid insertion, before and after an *in vivo* immune provocation. For this current analysis, we have included baseline (i.e. unprovoked) data at both the lumbar and deltoid sites and treated ‘anatomical site’ as repeated observations within participants (see *Statistical Analysis* section for more details).

### Participants, screening, and enrolment

To recruit participants with a varied range of childhood adversity, volunteers completed the 28-item Childhood Trauma Questionnaire-Short form (CTQ-SF) [6]. Using the REDCap electronic data capture tool hosted at the University of Cape Town [25, 26], pain-free adult volunteers (≥18 and ≤65 years old) were screened (eligibility details in Table 1) and enrolled into approximately equally sized groups to cover 1) minimal childhood adversity (control) (CTQ-SF score 25 – 36), 2) moderate childhood adversity (CTQ-SF score 37 – 67), and 3) severe childhood adversity (CTQ-SF score > 67) [5]. Participants were enrolled using a ‘first to qualify and participate’ approach. Group allocation was used only to recruit a sample with a varied range of childhood adversity. All participants underwent the same procedure.

**Table 1:** Eligibility criteria.

| Exclusion criteria | Safety risk |  |  | Confounding risk |  |
| --- | --- | --- | --- | --- | --- |
|  | General | HFS | Influenza vaccine | Psychophysical tests | Stimulated cytokines |
| Not fluent in English | X |  |  |  |  |
| Incompetence to consent and participate, e.g. acute psychosis or high suicide risk | X |  |  |  |  |
| Pregnancy |  | X |  |  | X |
| Electrical implants (e.g. pacemaker) |  | X |  |  |  |
| Metal implants in the area receiving the HFS |  | X |  |  |  |
| Tattoos in the area receiving the HFS |  | X |  |  |  |
| Any visible injury or open wounds in the area receiving the HFS |  | X |  |  |  |
| Cardiovascular disorders |  | X |  |  |  |
| Known history of allergic reactions to vaccinations |  |  | X |  |  |
| Already received current season's influenza vaccination |  |  | X |  | X |
| Chronic pain (pain on most days for the past three months) |  |  |  | X | X |
| Diabetes mellitus |  | X |  |  | X |
| Peripheral vascular disease |  | X |  | X |  |
| Sensory impairment of areas to undergo psychophysical testing |  | X |  | X |  |
| Use of medication that could alter skin |  |  |  | X | X |
| sensitivity (e.g. analgesic medication, topical medical creams in areas to undergo psychophysical testing) |  |  |  |  |  |
| Medication used to alter immune function (e.g. NSAIDs, steroids) |  |  |  |  | X |
| Smoking habit |  |  |  |  | X |
| Febrile illness in the past 4 weeks |  |  |  |  | X |
| Reasons for each criterion are specified using crosses in the applicable column. The criteria linked to possible safety risks of the influenza vaccine and confounding risks of the stimulated cytokines (both shown in grey) are relevant only for the primary study [3] and not to this current analysis. HFS: high-frequency electrical stimulation; NSAIDs: non-steroidal anti-inflammatory drugs. |  |  |  |  |  |

### Experimental tests

#### Induction of secondary hyperalgesia

Participants received high-frequency electrical stimulation at one forearm to induce secondary hyperalgesia. In the primary study, peripheral blood was drawn from one forearm; therefore, the high-frequency electrical stimulation was performed on the opposite arm to the blood draw to avoid stimulating an already sensitised area. The high-frequency electrical stimulation was delivered using a constant current stimulation system (DS7A, Digitimer Limited, Hertfordshire, UK) to one pair of specialised surface electrodes on the test forearm. The electrodes consist of a circular cathode (∼2 cm diameter) comprising 10 blunt steel pins arranged in a circle and applied to the anterior forearm, and a large, flexible fabric anode (∼5 cm width) placed around the upper arm, closely resembling the setup used by Henrich, Magerl [27], Klein, Magerl [34], Klein, Stahn [35]. The intensity of the high-frequency electrical stimulation was ten times the current of the individual’s detection threshold, which was determined using an adaptive staircase method, as previously described [3]. The high-frequency electrical stimulation consisted of five one-second trains, using a two-millisecond pulse width at a 100 Hz frequency, with a nine-second break between trains.

### Outcomes and measures

#### Sensation and Pain Ratings Scale

Participants provided stimulus ratings using the Sensation and Pain Ratings Scale (SPARS) [41]. The SPARS ranges from -50 (“no sensation”) to +50 (“most intense pain you can imagine”), with 0 representing “the point at which what you feel transitions to pain”.

Therefore, the SPARS comprehensively captures a range of ratings to both non-painful and painful stimuli.

#### Surface area of secondary hyperalgesia

The surface area (cm^2^) of secondary hyperalgesia was assessed along eight radial lines using a 128 mN von Frey filament (MARSTOCK, Schriesheim, Germany). We chose to use a 128 mN von Frey filament to map the area because our previous systematic review identified 128 mN as the most commonly used weight for mapping surface area of secondary skin hyperalgesia [4]. Dots were drawn 1 centimetre apart along each line, with the induction site at the central point of the radial lines. The assessor applied a mechanical punctate stimulus for approximately one second at each dot along each radial line, starting from the outside and moving towards the induction site at the centre of the radial lines. The participant was instructed to state when they feel a distinct increase in sensitivity to the mechanical punctate stimulation; this is typically perceived as an obvious increase. This point of increase was marked as the border of the surface area of SH. Surface area was mapped at 30, 45, and 60 minutes after the high-frequency electrical stimulation induction. We included the area data from each of the three time points in the statistical analysis, such that each participant contributed three surface area datapoints in total.

#### Magnitude of secondary hyperalgesia

Ratings to mechanical punctate stimulation were assessed using the SPARS [41] and two punctate “pinprick” stimulators that exerted forces of 128 mN and 256 mN (MRC Systems, Heidelberg, Germany) at baseline and at 35, 50, and 65 minutes after the induction. The two punctate “pinprick” stimulators were each applied three times approximately one centimetre adjacent to the cathode electrode. Participants provided an average rating using the SPARS for the three stimulations. We calculated the magnitude of secondary hyperalgesia by subtracting the baseline rating from the follow-up rating at each time point for each pinprick stimulator. Therefore, we included six ratings in total in the statistical analysis: one rating for each pinprick weight from each of the three time points.

#### Temporal summation

Temporal summation was assessed in the lumbar region (2 cm lateral to the second lumbar vertebra) and at the deltoid insertion. We calculated temporal summation by subtracting the SPARS rating of a single stimulation from the SPARS rating of the final of 16 stimulations at 60 Hz using a 256 mN Von Frey filament [1] to yield one measurement of temporal summation per participant per test site (i.e. lumbar and deltoid).

#### Conditioned pain modulation

Conditioned pain modulation (CPM) was also assessed in the lumbar region and at the deltoid insertion approximately 10 minutes after the assessments of temporal summation. The test stimulus was pressure pain threshold (N), and the conditioning stimulus was immersion of the contralateral hand in cold water of approximately 3 to 5 C. Pressure pain threshold was assessed with a hand-held algometer, with pressure increasing by approximately 5 N per second until report of first pain. Then, participants immersed their contralateral hand in circulating cold water until pain in the immersed hand reached 20 on the SPARS, representing “moderately painful” [41], at which time pressure pain threshold was reassessed while the contralateral hand remained immersed. Conditioned pain modulation was estimated by subtracting the pressure pain threshold before immersion from the pressure pain threshold during cold water immersion to yield one measurement of CPM per participant per test site.

#### Potential confounding factors

Candidate confounders were selected using a four-pronged approach: creating a directed acyclic graph, consulting with experts in the field, reviewing literature, and considering the feasibility of assessments [3] (Supplementary file; Section 2). The final candidate confounders prioritised for assessment were recent stress (Perceived Stress Scale [13]), self-reported diagnosis of major depressive disorder, acute illness(es) including COVID-19 infection within the preceding 6 months, chronic illness(es), and sleep quality and quantity (Pittsburgh Sleep Quality Index [11]). Additionally, the mean SPARS ratings of the five high- frequency electrical stimulation induction trains and the induction current were included as potential confounders for the outcomes of the surface area and magnitude of secondary hyperalgesia.

### Procedure

Figure 1 shows the full study procedure with outcomes relevant to this analysis displayed in colour. Participants were orientated to the procedure and gave written consent. Participants then underwent assessments of temporal summation and CPM and provided baseline SPARS ratings to mechanical stimulation. Thereafter, secondary hyperalgesia was induced using high-frequency electrical stimulation. The surface area of secondary hyperalgesia was mapped at 30, 45, and 60 minutes after the high-frequency electrical stimulation induction, and follow-up ratings to mechanical stimulation were obtained at 35, 50, and 65 minutes after the induction.

**Figure 1:**
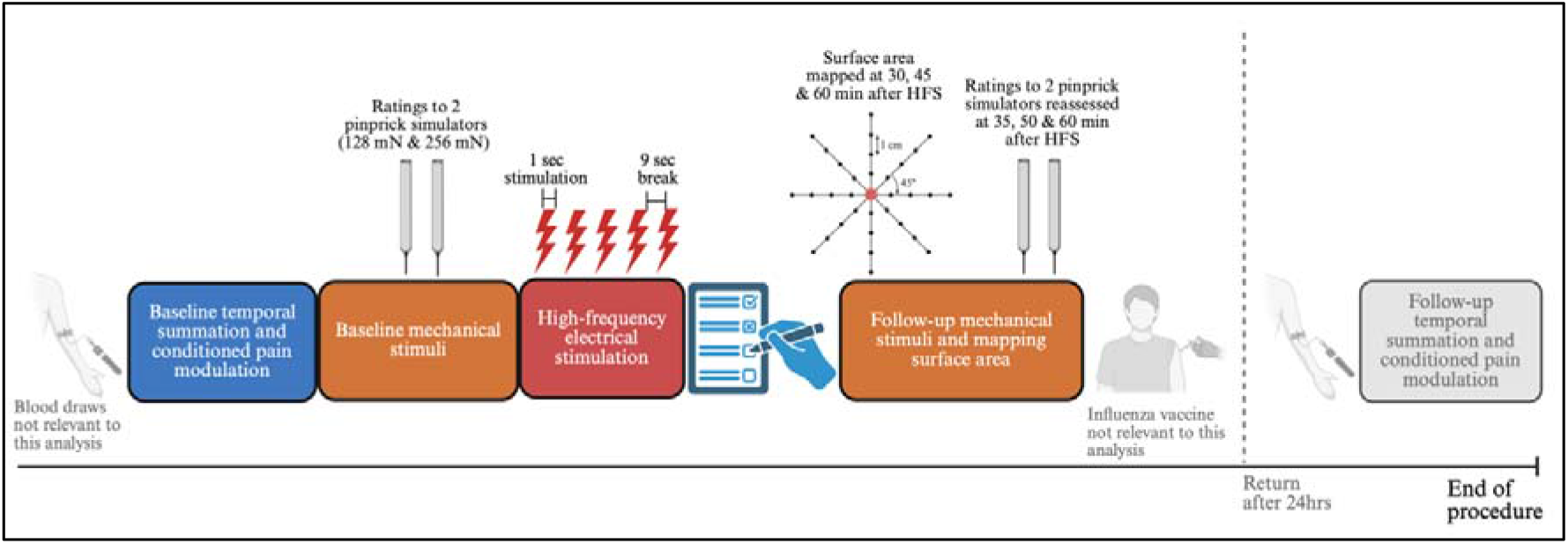
Study procedure [3], with outcomes relevant to this secondary analysis displayed in colour. First, participants underwent baseline assessments of temporal summation and CPM at the lumbar and deltoid sites. Second, participants gave ratings to two mechanical pinprick stimulators exerting forces of 128 mN and 256 mN each. Third, participants received the high-frequency electrical stimulation at one forearm. Fourth, participants completed questionnaires to assess potential confounders. Last, the surface area of secondary hyperalgesia was mapped using the 8-radial-lines approach at 30, 45, and 60 minutes after the induction, and the ratings to mechanical pinprick stimulation were re-assessed at 35, 50, and 65 minutes after the induction. As displayed in grey, blood draws, the influenza vaccination, and follow-up assessments of temporal summation and CPM were not relevant to this secondary analysis. Figure created in BioRender: https://BioRender.com/q32i088.

### Statistical analysis plan

The sample size of 96 participants was determined by the primary study [3]; therefore, it was fixed for this secondary analysis. Notably, we excluded data from one participant due to sex hormone therapy, which could interfere with the current analysis. Therefore, we had a total sample size of N = 95 participants.

We conducted an approximate sensitivity power analysis using G*Power version 3.1 [16] to determine the minimum incremental effect size that could be detected with the available sample. The fully adjusted regression models differed between outcomes; therefore, we performed separate sensitivity analyses. The models for the surface area and magnitude of secondary hyperalgesia included the exposure of interest and 10 prespecified covariates (11 predictors in total), whereas the models for conditioned pain modulation and temporal summation included the exposure of interest and eight prespecified covariates (9 predictors in total). Assuming a two-sided significance level of 0.05, 80% power, and a sample size of 95 participants, the minimum detectable incremental effect sizes were 0.085 for both model specifications, indicating that this analysis had sensitivity to detect effects of approximately this magnitude or larger.

Data visualisation was undertaken before formal statistical modelling to examine distributions, identify potential outliers, assess model assumptions, and explore the relationships between childhood adversity and experimental tests of central pain modulation. Four pre-specified primary hypotheses were tested: that CTQ-SF total score would be associated with

1. larger surface areas of induced secondary hyperalgesia,
2. increased magnitudes of induced secondary hyperalgesia,
3. greater temporal summation, and
4. reduced CPM.

To account for multiple testing across these four primary hypotheses, the false discovery rate was controlled using the Benjamini-Hochberg procedure. Adjusted *q*-values are reported alongside the corresponding *p*-values. The *p*-values reflect the statistical evidence for each individual hypothesis before correction for multiple testing, whereas the *q*-values account for the testing of all four primary hypotheses and should therefore be used to determine statistical significance after adjustment for multiple comparisons. A *q*-value < 0.05 was considered statistically significant.

We also hypothesised that the relationship between CTQ-SF total score and each of the experimental pain outcomes would differ between sexes. Therefore, we included sex as an interaction term in the regression models. For each primary outcome, three progressively adjusted regression models were fitted to evaluate the same underlying hypothesis.

Model 1: We investigated the main effect of childhood adversity on each experimental pain outcome using an unadjusted regression model.

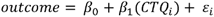

Model 2: We added sex as an interaction term to assess whether the association between childhood adversity and the experimental pain outcome differed between sex.

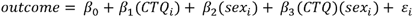

Model 3: Finally, we adjusted for relevant covariates that could influence the experimental pain outcomes.

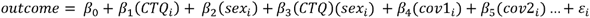

1 2 … Hypotheses 1 and 2 were tested using linear mixed-effects regression models, with CTQ-SF total score as the independent variable and the surface area or magnitude of secondary hyperalgesia as the dependent variable. Participant was modelled as a random effect for both outcomes, with pinprick weight (128 mN and 256 mN) additionally included as a random effect nested within participant for the magnitude of secondary hyperalgesia.

Hypotheses 3 and 4 were tested using linear mixed-effects regression models, with CTQ-SF total score as the independent variable and temporal summation or CPM as the dependent variable. Measurement site (lumbar and deltoid) was modelled as repeated observations within participants. To determine whether the association between CTQ-SF total score and temporal summation/CPM differed by anatomical site, a CTQ-SF × site interaction term was included. Where there was no evidence of interaction, the overall association across sites was reported; where an interaction was present, site-specific estimates were reported as secondary analyses.

Data were cleaned and analysed using R (packages: readr [68], tidyverse [66], magrittr [42], ggplot2 [65], dplyr [67], lme4 [2], patchwork [46], sjPlot [39], MASS [64], lmerTest [37], robustlmm [36], pbapply [55], and parameters [40]) in RStudio (version 2024.4.1, Build 748) [51].

#### Exploratory analysis

We investigated the relationship between recent stress and each of the four experimental pain outcomes. Recent stress was operationalised by the total score on the Perceived Stress Scale (PSS) (possible range: 0 – 40) [13]. We used the same approach of three progressively adjusted regression models to evaluate the same underlying hypothesis as explained above, except that the PSS served as the independent variable. We hypothesised that the score on the PSS would be positively associated with the surface area and magnitude of secondary hyperalgesia and temporal summation, and negatively associated with CPM. Given that this analysis was exploratory, we did not correct for multiple testing.

#### Assessment of model fit

Assessments of model fit considered the assumptions of (1) linearity, (2) homoscedasticity, (3) normally distributed residuals, and (4) no influential observations. A model was deemed a poor fit to the data if any assumption was violated. If the assumption of no influential observations was violated, we planned to proceed with a robust linear regression because conventional (i.e. ordinary least squares) models are sensitive to outliers and may provide inaccurate estimations of the β coefficient in the presence of influential observations [63]. Robust regression models, as implemented using the robustlmm package in R, are less influenced by extreme values than the conventional linear regression model [36]. While robust models are designed to reduce the influence of outliers, they do not address violations of linearity and homoscedasticity. To address these violations, we planned to bootstrap the confidence intervals of the β coefficients from the model (robust or conventional). Bootstrapping assumes neither linearity nor homoscedasticity; thus, it reduces the risk of type 1 error and provides a more accurate estimate of the 95% confidence intervals when homoscedasticity is violated [50].

## Results

Ninety-five participants (61 females; median (range) age: 23 (18 – 65) years old) were included in this analysis. There were complete datasets for all outcomes except that one participant was missing temporal summation data at the lumbar site, due to a technical problem. This participant was excluded from only the analysis of temporal summation at the lumbar site. Plots of model assumptions are presented in the Supplementary File, Section 3, and reported here only if model assumptions were violated.

Descriptive statistics are presented in Table 2. Compared to females, male participants were slightly younger and had a higher electrical detection threshold and therefore a higher current for the high-frequency electrical stimulation (median (IQR): 0.19 (0.16 – 0.25) mA vs 0.17 (0.13 – 0.20) mA, *p* = 0.02). Male participants also had a larger CPM effect at both the lumbar and deltoid sites than females (median (IQR) effect on PPT at lumbar: increase of 22.35 N (17.19 – 32.64) for males vs 17.20 N (11.15 – 27.20) for females, *p* = 0.02; median (IQR) effect at deltoid: 14.85 N (8.44 – 21.43) for males vs 9.55 N (5.45 – 15.55) for females, *p* = 0.02). There were no statistically significant differences between sexes for CTQ-SF total score and subscales scores, the surface area and magnitude of secondary hyperalgesia, temporal summation, or recent stress (Table 2).

**Table 2:** Summary of descriptive statistics for the full sample and stratified by sex.

| Characteristics | Full sample<br>N = 95 | Females<br>n = 61 | Males<br>n = 34 | p-value for<br>between-sex<br>differences |
| --- | --- | --- | --- | --- |
| Age (years) | 23 (21– 31) | 24 (21 – 35) | 21 (21 – 26) | **0.036** |
| Adverse childhood<br>experiences (CTQ-SF)* | 49 (33 – 74) | 48 (32 – 75) | 49 (38 – 63) | 0.55 |
| Subscale: physical abuse<sup>†</sup> | 8.0 (5.0 – 14.5) | 8.0 (5.0 – 14.0) | 7.0 (6.0 – 14.0) | 0.99 |
| Subscale: emotional abuse<sup>†</sup> | 10.0 (7.0 – 19.0) | 13.0 (7.0 – 20.0) | 9.0 (8.0 – 15.0) | 0.18 |
| Subscale: sexual abuse<sup>†</sup> | 5.0 (5.0 – 12.5) | 5.0 (5.0 – 14.0) | 5.0 (5.0 – 10.8) | 0.26 |
| Subscale: physical neglect<sup>†</sup> | 8.0 (5.0 – 13.0) | 8.0 (5.0 – 12.0) | 8.5 (5.3 – 13.8) | 0.73 |
| Subscale: emotional neglect<sup>†</sup> | 13.0 (7.0 – 19.0) | 13.0 (7.0 – 19.0) | 12.5 (7.3 – 18.0) | 0.96 |
| **Current used for HFS (mA)** | 0.17 (0.13 – 0.23) | 0.17 (0.13 – 0.20) | 0.19 (0.16 – 0.25) | **0.019** |
| **Mean SPARS ratings to 5 HFS induction trains<sup>‡</sup>** | 32.92 (26.03 – 42.83) | 34.38 (26.84 – 44.16) | 30.25 (23.97 – 38.48) | 0.064 |
| **Surface area of secondary hyperalgesia (cm<sup>2</sup>)** |  |  |  |  |
| 30 min after HFS | 20.15 (5.48 – 34.12) | 18.39 (3.54 – 33.59) | 22.10 (9.28 – 34.65) | 0.95 |
| 45 min after HFS | 15.56 (6.72 – 32.53) | 14.85 (5.30 – 32.53) | 20.33 (9.28 – 32.35) | 0.89 |
| 60 min after HFS | 15.56 (3.36 – 28.46) | 15.91 (3.54 – 28.28) | 14.67 (2.21 – 29.08) | 0.52 |
| **Magnitude of secondary hyperalgesia (change in SPARS ratings, follow-up minus baseline)** |  |  |  |  |
| 35 min after HFS | 1.87 (-2.59 – 9.78) | 2.16 (-4.29 – 9.40) | 1.71 (-1.09 – 10.13) | 0.34 |
| 50 min after HFS | 4.13 (-1.74 – 11.74) | 3.86 (-2.43 – 11.74) | 4.70 (-0.29 – 11.25) | 0.97 |
| 65 min after HFS | 4.48 (-1.08 – 13.85) | 4.15 (-1.08 – 14.56) | 4.75 (-0.92 – 11.15) | 0.91 |
| **Temporal summation (change in SPARS ratings, 16<sup>th</sup> minus 1st)** |  |  |  |  |
| Lumbar test site | 6.39 (1.27 – 19.27) | 6.87 (1.39 – 21.27) | 5.64 (1.05 – 18.59) | 0.20 |
| Deltoid test site | 7.46 (0.99 – 17.98) | 7.46 (1.67 – 17.14) | 7.78 (0.81 – 18.00) | 0.96 |
| **Conditioned pain modulation (change in pressure pain threshold, N)** |  |  |  |  |
| Lumbar test site | 20.80 (12.48 – 29.58) | 17.20 (11.15 – 27.20) | 22.35 (17.19 – 32.64) | **0.016** |
| Deltoid test site | 11.45 (5.93 – 18.63) | 9.55 (5.45 – 15.55) | 14.85 (8.44 – 21.43) | **0.021** |
| **Recent stress (PSS)<sup>§</sup>** | 17.00 (12.50 – 22.50) | 18.00 (14.00 – 23.00) | 16.50 (12.00 – 19.00) | 0.14 |

Data are presented as median (IQR). Statistical test for all variables was an ANOVA. P-values are rounded to two significant decimal points.
\*Possible total score range for the CTQ-SF: 25 to 125.
<sup>†</sup>Possible total score range for each subscale of the CTQ-SF: 5 to 25.
<sup>‡</sup>Possible range for the SPARS is -50 to +50.
<sup>§</sup>Possible range for the PSS is 0 to 40.
CTQ-SF = Childhood Trauma Questionnaire-short form; HFS = high-frequency electrical stimulation; PSS = Perceived Stress Scale; SPARS = Sensation and Pain Rating Scale.

Childhood adversity was positively associated with the surface area of induced secondary hyperalgesia.

In the conventional mixed-effects linear regression model to investigate the relationship between CTQ-SF total score and the surface area of secondary hyperalgesia, the assumptions of linearity, homoscedasticity, and absence of influential observations were violated (Supplementary file: Section 3a, Fig S1). Therefore, we proceeded with a robust mixed-effects linear regression to reduce the influence of extreme values and bootstrapped the 95% confidence intervals of the effect estimates generated by the robust, covariate-adjusted mixed-effects model.

The unadjusted robust model found a positive association between CTQ-SF total score and the surface area of secondary hyperalgesia (β=0.17 [95%CI: 0.02; 0.32], *p* = 0.025) (Fig 2 and Table 3). When sex was included as an interaction term, both the unadjusted and covariate-adjusted robust models also found a positive association between CTQ-SF total score and the surface area of secondary hyperalgesia (effect of CTQ-SF on area, unadjusted: β=0.36 [95%CI: 0.06; 0.65], *p* = 0.019; adjusted: β=0.32 [95%CI: 0.01; 0.62], *p* = 0.043) (Table 3). Although visual inspection of the predicted surface area of secondary hyperalgesia suggests a potential interaction between CTQ-SF total score and sex, as indicated by the crossing lines and a stronger association between CTQ-SF total score and the surface area of secondary hyperalgesia in males than in females (Fig 2b), the robust model showed no evidence of a statistically significant effect of sex, nor an interaction between CTQ-SF total score and sex (Table 3).

**Figure 2:**
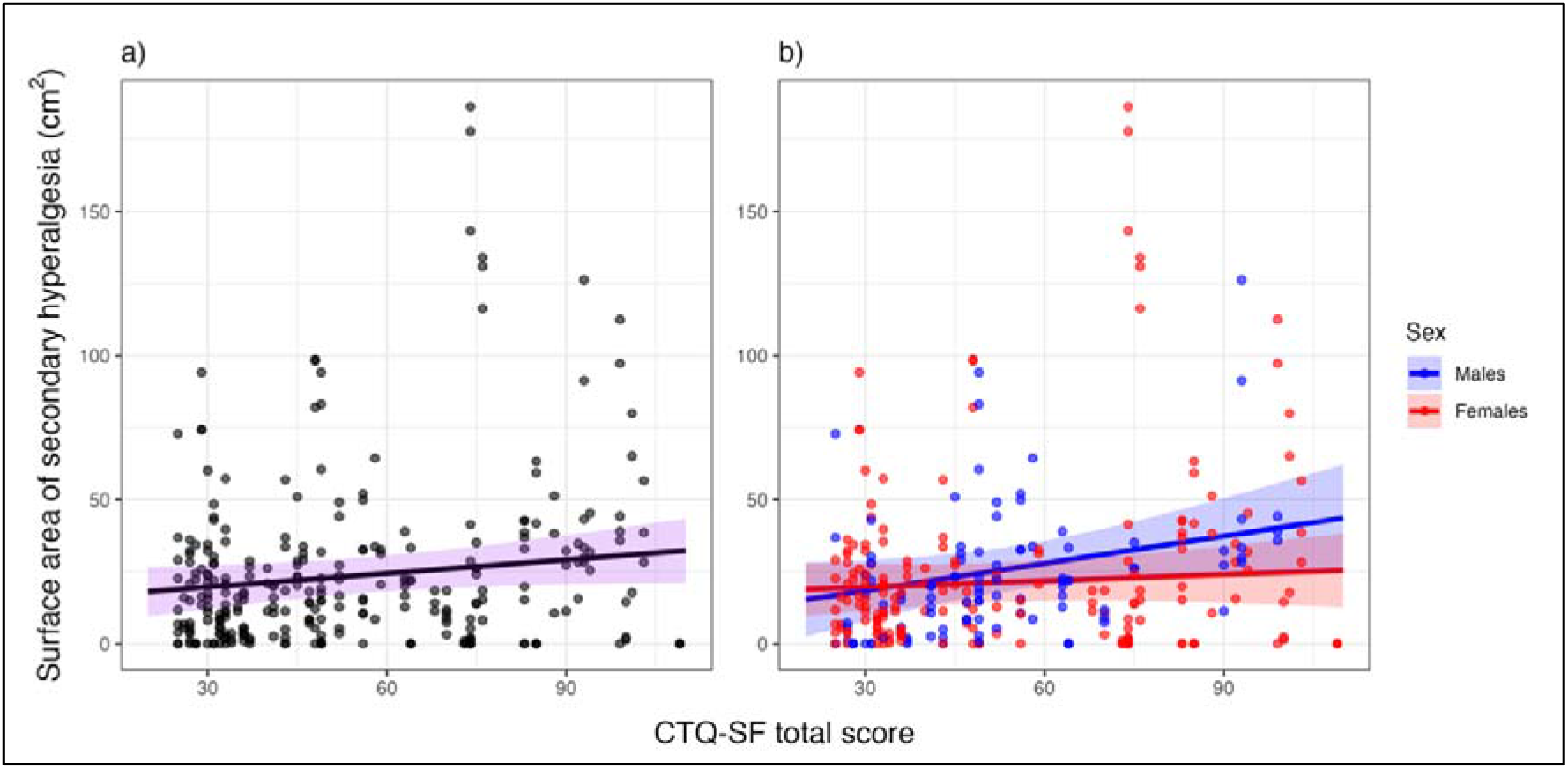
Plot of the association between the Childhood Trauma Questionnaire-Short Form (CTQ-SF) total score and the surface area of secondary hyperalgesia. Plots show the observed values (dots) and the predicted regression line and 95% confidence interval (ribbon) for the covariate-adjusted model for the full cohort (plot a) and with sex as an interaction term (plot b).

**Table 3:** Details of models of the relationship between childhood adversity (CTQ-SF total score) and the surface area of induced secondary hyperalgesia, with and without covariates.

|  | Unadjusted model of main effect |  |  | Unadjusted model with sex interaction term |  |  | Adjusted model with sex interaction term |  |  | Bootstrapped 95%CI of the adjusted model with sex interaction term |
| --- | --- | --- | --- | --- | --- | --- | --- | --- | --- | --- |
| Predictors | Estimates | 95% CI | p | Estimates | 95% CI | p | Estimates | 95% CI | p |  |
| Intercept | 10.50 | 1.65; 19.36 | **0.020** | 2.03 | -14.56; 18.61 | 0.81 | 5.97 | -19.15; 31.09 | 0.64 | -18.58; 28.47 |
| CTQ-SF score | 0.17 | 0.02; 0.32 | **0.025** | 0.36 | 0.06; 0.65 | **0.019** | 0.32 | 0.01; 0.62 | **0.043** | 0.09; 0.63 |
| Sex (female) |  |  |  | 11.63 | -7.95; 31.22 | 0.24 | 8.49 | -11.26; 28.24 | 0.40 | -7.37; 27.84 |
| CTQ-SF score x sex (female) |  |  |  | -0.26 | -0.60; 0.09 | 0.14 | -0.24 | -0.59; 0.10 | 0.17 | -0.60; 0.09 |
| Age |  |  |  |  |  |  | 0.46 | 0.05; 0.87 | **0.028** | 0.08; 0.93 |
| Mean rating of HFS induction |  |  |  |  |  |  | -0.06 | -0.38; 0.26 | 0.72 | -0.37; 0.22 |
| HFS current intensity |  |  |  |  |  |  | -37.13 | -79.98; 5.72 | 0.09 | -74.54; -11.23 |
| PSS score |  |  |  |  |  |  | 0.06 | -0.59; 0.71 | 0.86 | -0.60; 0.68 |
| PSQI score |  |  |  |  |  |  | -0.25 | -1.36; 0.86 | 0.66 | -1.41; 0.79 |
| Diagnosis of major depressive disorder |  |  |  |  |  |  | 1.17 | -12.65; 15.00 | 0.87 | -12.90; 17.08 |
| Recent COVID-19 infection |  |  |  |  |  |  | 5.26 | -9.79; 20.30 | 0.49 | -5.81; 20.43 |
| Recent acute illness |  |  |  |  |  |  | -4.76 | -12.81; 3.30 | 0.25 | -12.31; 2.74 |
| Diagnosis of chronic illness |  |  |  |  |  |  | -6.01 | -18.35; 6.33 | 0.34 | -20.85; 7.80 |
| Random effects |  |  |  |  |  |  |  |  |  |  |
| $\sigma^2$ | 167.17 | | | 66.17 | | | 65.28 | | | |
| $\tau_{00}$ | 647.29<sub>study_id</sub> | | | 258.75<sub>study_id</sub> | | | 250.20<sub>study_id</sub> | | | |
| ICC | 0.79 |  |  | 0.80 |  |  | 0.79 |  |  |  |
| N | 95<sub>study_id</sub> |  |  | 95<sub>study_id</sub> |  |  | 95<sub>study_id</sub> |  |  |  |
| Observations | 285 |  |  | 285 |  |  | 285 |  |  |  |
| Marginal R<sup>2</sup> / Conditional R<sup>2</sup> | 0.043/0.804 |  |  | 0.063/0.809 |  |  | 0.151/0.824 |  |  |  |
| P-values are rounded to two significant decimal points. |  |  |  |  |  |  |  |  |  |  |
| CTQ-SF = Childhood Trauma Questionnaire-short form; HFS = high-frequency electrical stimulation; PSS = Perceived Stress Scale; PSQI = Pittsburgh Sleep Quality Index. |  |  |  |  |  |  |  |  |  |  |

Bootstrapping produced little change in the 95% CI for the effect of CTQ-SF total score on the surface area of SH, using the robust, covariate-adjusted model (Table 3), and the model maintained that CTQ-SF total score was positively associated with the surface area of SH. The bootstrapped 95% CI translates to an effect size ranging from small (0.09) to large (0.63), suggesting low precision in the effect estimate (Table 3). The bootstrapped 95% CIs for sex and for the interaction between CTQ-SF total score and sex both crossed zero.

Therefore, we cannot reject the null hypotheses of no effect of sex and no effect of the interaction between CTQ-SF score and sex at the 5% threshold for significance (Table 3).

There was no evidence of an association between childhood adversity and the magnitude of induced secondary hyperalgesia.

In the conventional mixed-effects linear regression model to investigate the relationship between CTQ-SF total score and the magnitude of secondary hyperalgesia, the assumptions of linearity, homoscedasticity, and no influential observations were violated (Supplementary file: Section 3b, Fig S2). Therefore, we proceeded with a robust mixed-effects linear regression and bootstrapped the 95% CIs of the robust, covariate-adjusted mixed-effects model.

The unadjusted robust model found no evidence of an association between CTQ-SF total score and the magnitude of secondary hyperalgesia (β= 0.01 [95%CI: -0.08; 0.10], *p* = 0.81; Fig 3 & Table 4). When sex was included as an interaction term, both the unadjusted (β= - 0.08 [95%CI: -0.26; 0.10], *p* = 0.37) and the covariate-adjusted robust models (β= -0.10 [95%CI: -0.28; 0.08], *p* = 0.28) found no evidence of an association between CTQ-SF total score and the magnitude of secondary hyperalgesia, and no evidence of an effect of sex, nor an interaction between CTQ-SF total score and sex (Table 4).

**Figure 3:**
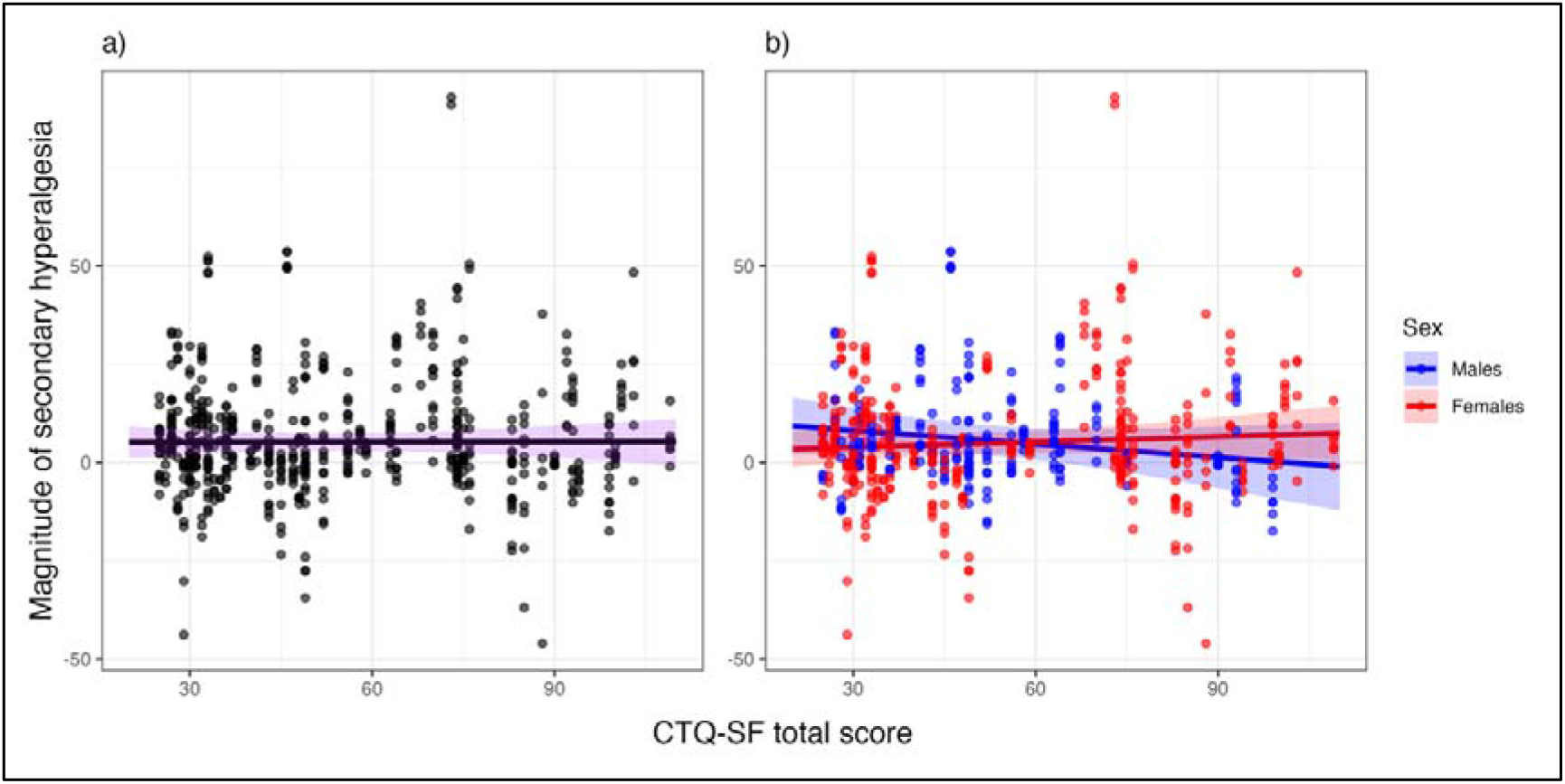
Plot of the association between the Childhood Trauma Questionnaire-Short Form (CTQ-SF) total score and magnitude of secondary hyperalgesia. Plots show the observed values (dots) and the predicted regression line and 95% confidence interval (ribbon) for the covariate-adjusted model for the full cohort (plot a) and with sex as an interaction term (plot b). Y axes have been truncated but range from -100 to +100.

**Table 4:**
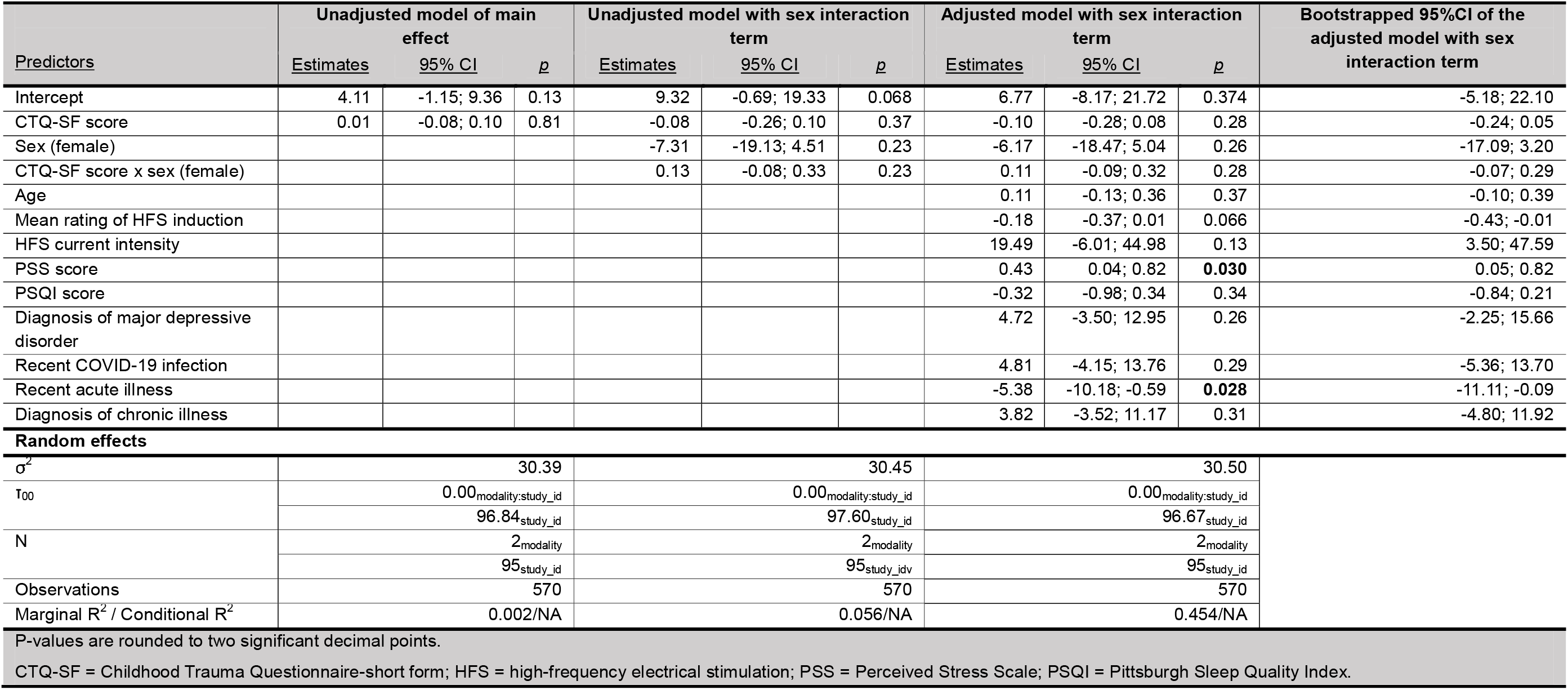
Summary of the relationship between childhood adversity (CTQ-SF total score) and magnitude of induced secondary hyperalgesia.

The bootstrapped 95% CI of the robust, covariate-adjusted robust model crosses zero (- 0.24; 0.05). Therefore, we cannot reject the null hypothesis of no effect of CTQ-SF total score on the magnitude of secondary hyperalgesia (Table 4). The bootstrapped 95% CIs for sex and for the interaction between CTQ-SF total score and sex both crossed zero.

Therefore, we cannot reject the null hypotheses of no effect of sex and no effect of the interaction between CTQ-SF score and sex at the 5% threshold for significance (Table 4).

There was no evidence of an association between childhood adversity and temporal summation.

On average, temporal summation was successfully induced at the sample level at both test sites (i.e. lumbar and deltoid): ratings of the final of 16 stimulation were higher than ratings of the first stimulation (Fig 4). On average, ratings increased by 10.09 units (95% CI: 7.65; 12.53, *p* <0.001; on the SPARS with range: -50 to +50) at the lumbar and 11.35 units (95% CI: 7.86; 14.39, *p* <0.001) at the deltoid site. Most of the ratings for both the first and the final of 16 stimulations were less than zero, indicating that they were in the “non-painful” range of the SPARS, despite using a 256 mN Von Frey filament as recommended by the DFNS Quantitative Sensory Testing manual [49]. At the lumbar site, 87 (of 94) of the first stimulations and 74 (of 94) of the final of 16 stimulations were rated as less than zero – indicating they were perceived as non-painful. At the deltoid site, 92 (of 95) of the first stimulations and 82 (of 95) of the final of 16 stimulations were rated as less than zero.

**Figure 4:**
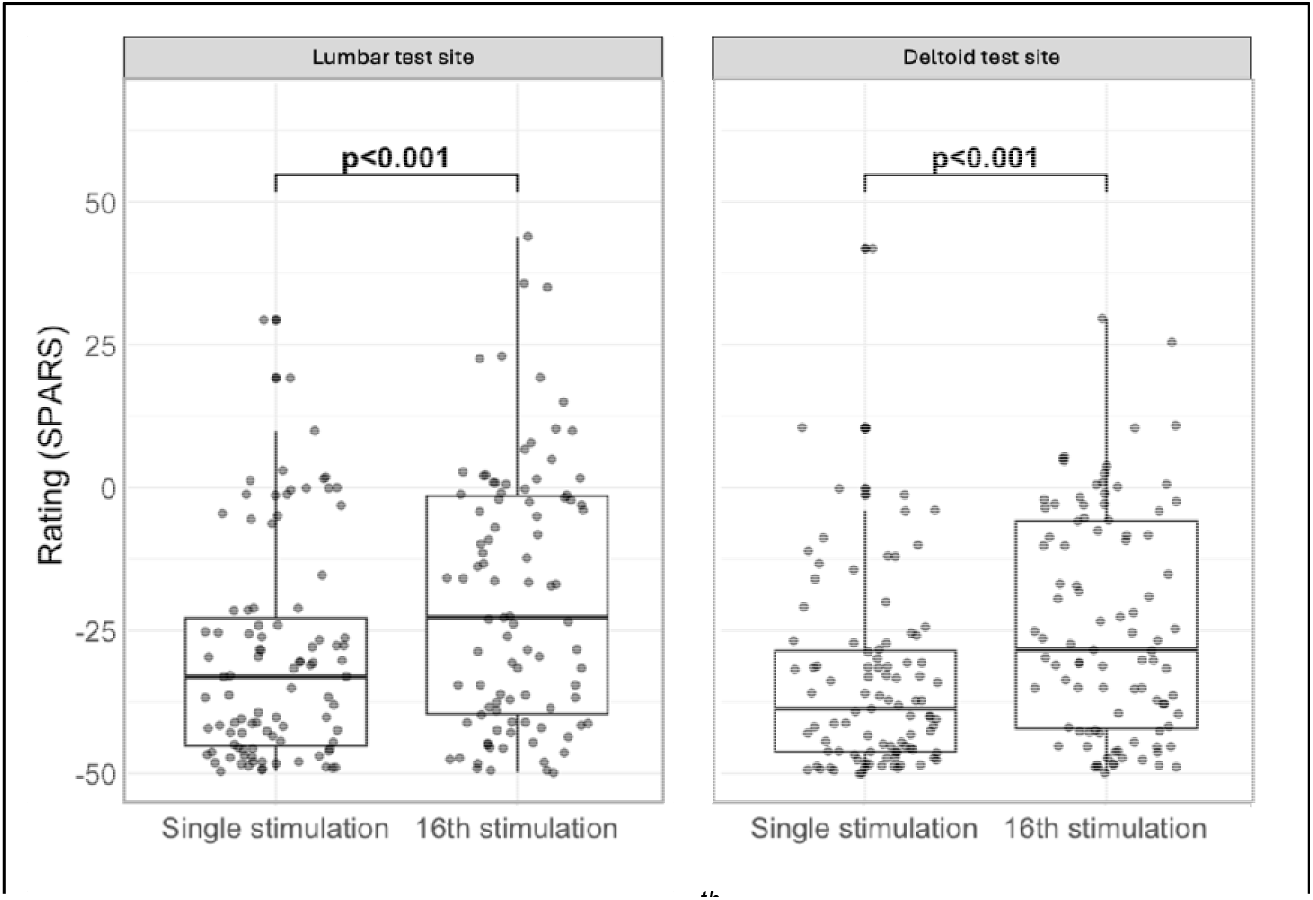
Boxplots of ratings to single and 16^th^ mechanical stimulation, faceted by test site.

In the unadjusted mixed-effects model accounting for repeated measurements and anatomical site, there was no evidence of an association between CTQ-SF total score and temporal summation (β = 0.04, 95% CI −0.06 to 0.14, p = 0.40; Fig 5 & Table 5). Temporal summation did not differ between the lumbar and deltoid sites (β = −1.2, 95% CI −4.7 to 2.3, p = 0.50; Table 5). There was no evidence that sex modified the association between CTQ-SF and temporal summation (CTQ-SF × sex interaction: β = 0.03, 95% CI −0.20 to 0.26, p = 0.80). After adjustment for covariates temporal summation remained non-significant (β = 0.03, 95% CI −0.17 to 0.23, p = 0.80; Table 5), and there was still no evidence of effect modification by sex (β = −0.02, 95% CI −0.25 to 0.21, p = 0.80). Temporal summation did not differ between the lumbar and deltoid sites after adjustment (β = −1.2, 95% CI −4.7 to 2.2, p = 0.50; Table 5).

**Figure 5:**
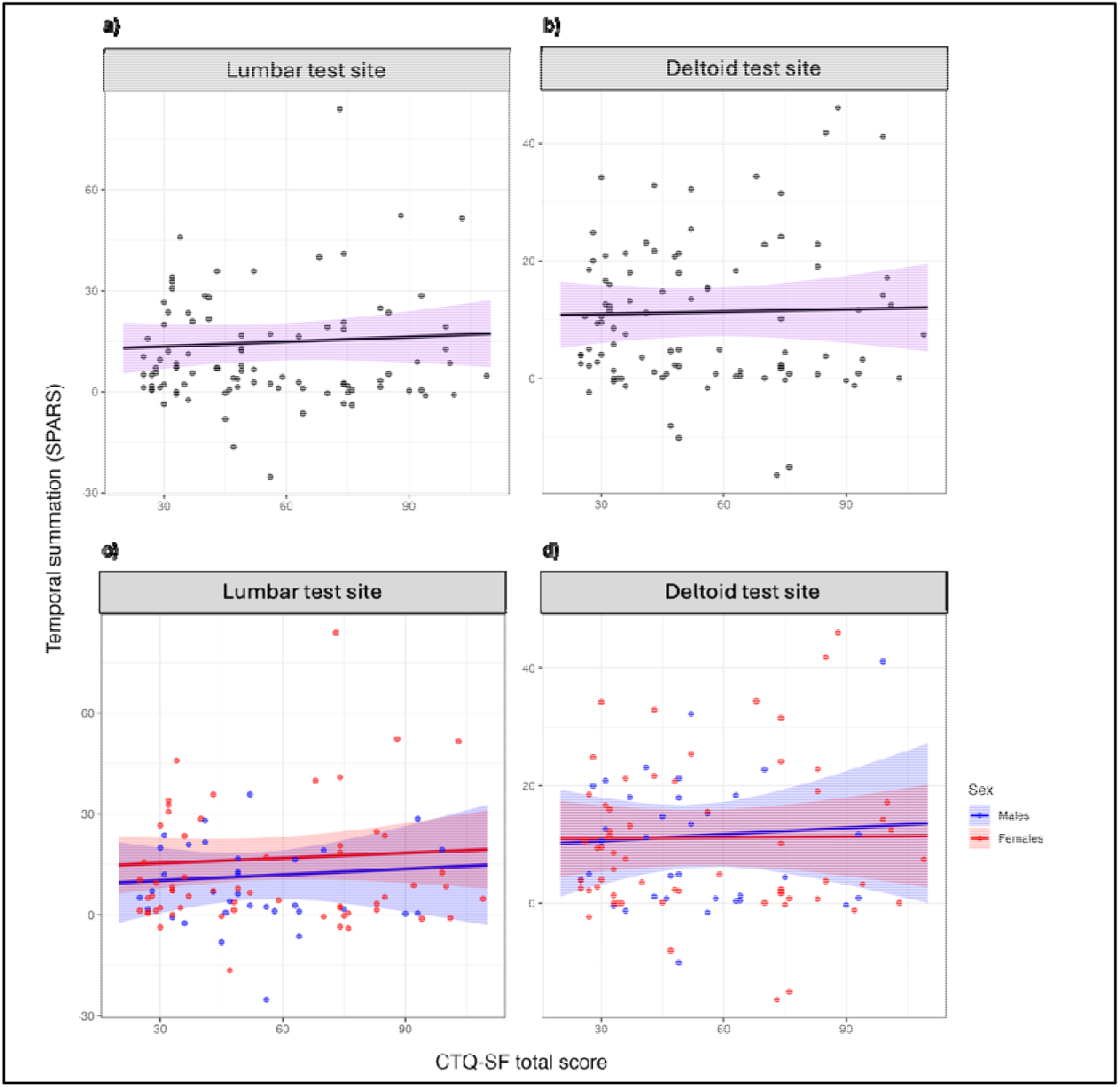
Plot of the association between the Childhood Trauma Questionnaire-Short Form (CTQ-SF) total score and temporal summation. Plots show the observed values (dots) and the predicted regression line and 95% confidence interval (ribbon) for the covariate-adjusted model for the full cohort (plots a and b) and with sex as an interaction term (plots c and d). Y axes have been truncated but range from -100 to +100

**Table 5:**
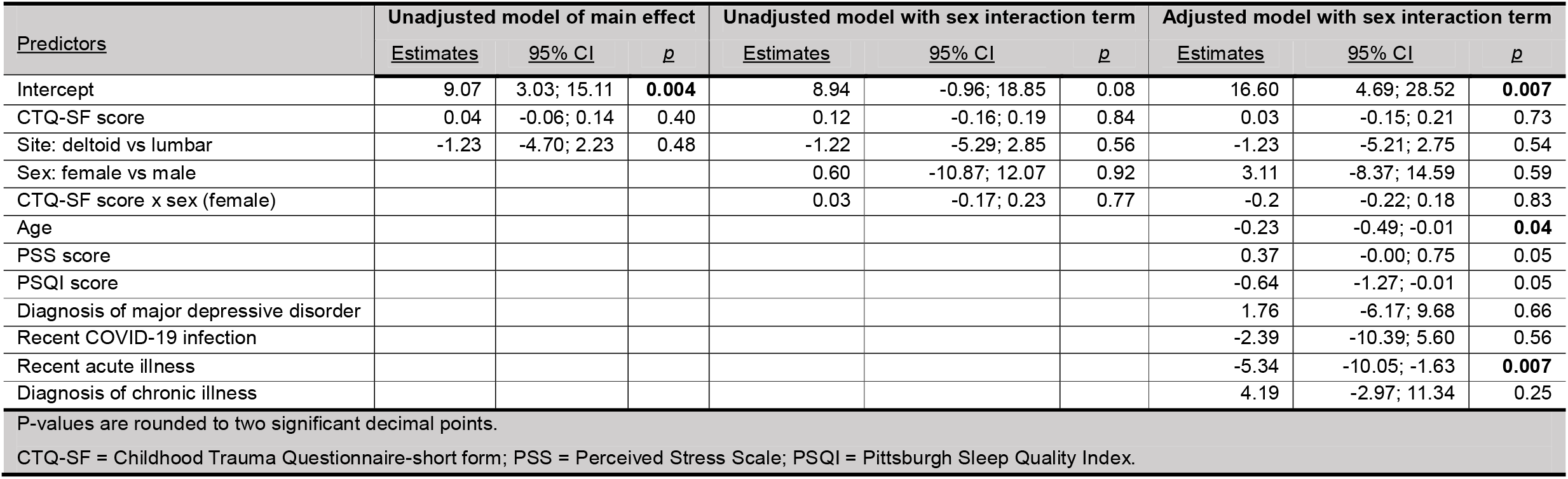
Summary of the relationship between childhood adversity (CTQ-SF total score) and temporal summation.

| Predictors | Unadjusted model of main effect |  |  | Unadjusted model with sex interaction term |  |  | Adjusted model with sex interaction term |  |  |
| --- | --- | --- | --- | --- | --- | --- | --- | --- | --- |
|  | Estimates | 95% CI | p | Estimates | 95% CI | p | Estimates | 95% CI | p |
| Intercept | 9.07 | 3.03; 15.11 | **0.004** | 8.94 | -0.96; 18.85 | 0.08 | 16.60 | 4.69; 28.52 | **0.007** |
| CTQ-SF score | 0.04 | -0.06; 0.14 | 0.40 | 0.12 | -0.16; 0.19 | 0.84 | 0.03 | -0.15; 0.21 | 0.73 |
| Site: deltoid vs lumbar | -1.23 | -4.70; 2.23 | 0.48 | -1.22 | -5.29; 2.85 | 0.56 | -1.23 | -5.21; 2.75 | 0.54 |
| Sex: female vs male |  |  |  | 0.60 | -10.87; 12.07 | 0.92 | 3.11 | -8.37; 14.59 | 0.59 |
| CTQ-SF score x sex (female) |  |  |  | 0.03 | -0.17; 0.23 | 0.77 | -0.2 | -0.22; 0.18 | 0.83 |
| Age |  |  |  |  |  |  | -0.23 | -0.49; -0.01 | **0.04** |
| PSS score |  |  |  |  |  |  | 0.37 | -0.00; 0.75 | 0.05 |
| PSQI score |  |  |  |  |  |  | -0.64 | -1.27; -0.01 | 0.05 |
| Diagnosis of major depressive disorder |  |  |  |  |  |  | 1.76 | -6.17; 9.68 | 0.66 |
| Recent COVID-19 infection |  |  |  |  |  |  | -2.39 | -10.39; 5.60 | 0.56 |
| Recent acute illness |  |  |  |  |  |  | -5.34 | -10.05; -1.63 | **0.007** |
| Diagnosis of chronic illness |  |  |  |  |  |  | 4.19 | -2.97; 11.34 | 0.25 |
| P-values are rounded to two significant decimal points. |  |  |  |  |  |  |  |  |  |
| CTQ-SF = Childhood Trauma Questionnaire-short form; PSS = Perceived Stress Scale; PSQI = Pittsburgh Sleep Quality Index. |  |  |  |  |  |  |  |  |  |

There was no evidence of an association between childhood adversity and conditioned pain modulation.

On average, CPM was successfully induced at the sample level at both test sites (i.e. lumbar and deltoid): pressure pain threshold was higher during the cold water immersion (i.e. conditioning stimulus) than before (Fig 6). On average, the cold water immersion increased pressure pain threshold by 21.10 N (95%CI: 18.09; 24.12, *p* <0.001) at the lumbar and by 13.04 (95%CI: 10.71; 15.36, *p* <0.001) at the deltoid site.

**Figure 6:**
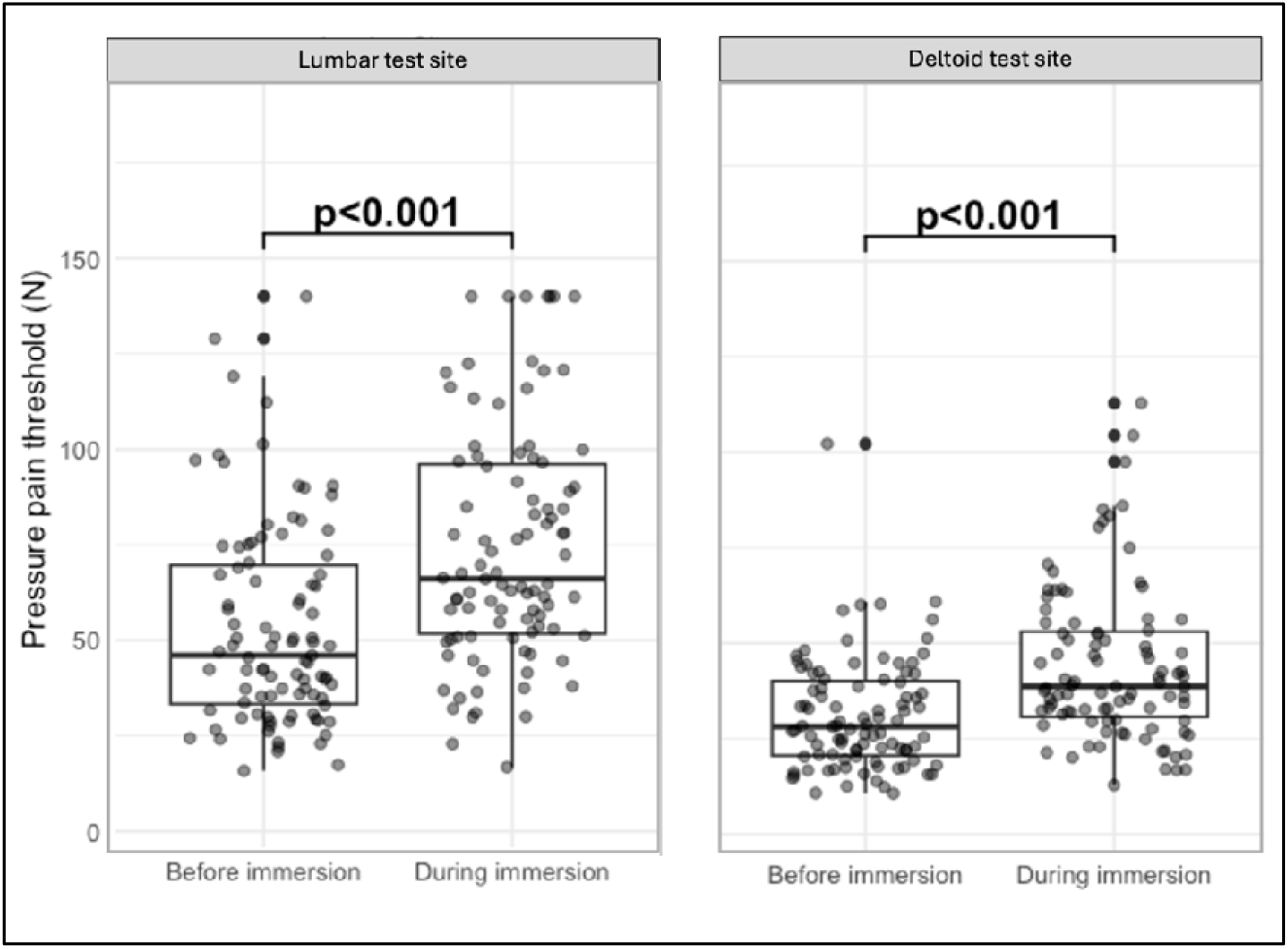
Boxplots of pressure pain threshold before and during cold water immersion, faceted by test site.

In the unadjusted mixed-effects model accounting for repeated measurements and anatomical site, there was no evidence of an association between CTQ-SF total score and CPM (β = 0.05, 95% CI −0.04 to 0.15, p = 0.30; Fig 7 & Table 6). There was no evidence that sex modified the association between CTQ-SF total score and CPM (CTQ-SF × sex interaction: β = −0.05, 95% CI −0.27 to 0.16, p = 0.60; Table 6). After adjustment for covariates, it remained that there was no evidence of an association between CTQ-SF total score and CPM (β = 0.11, 95% CI −0.08 to 0.30, p = 0.20; Table 6), and there was still no evidence of effect modification by sex (β = −0.05, 95% CI −0.26 to 0.17, p = 0.70; Table 6).

**Figure 7:**
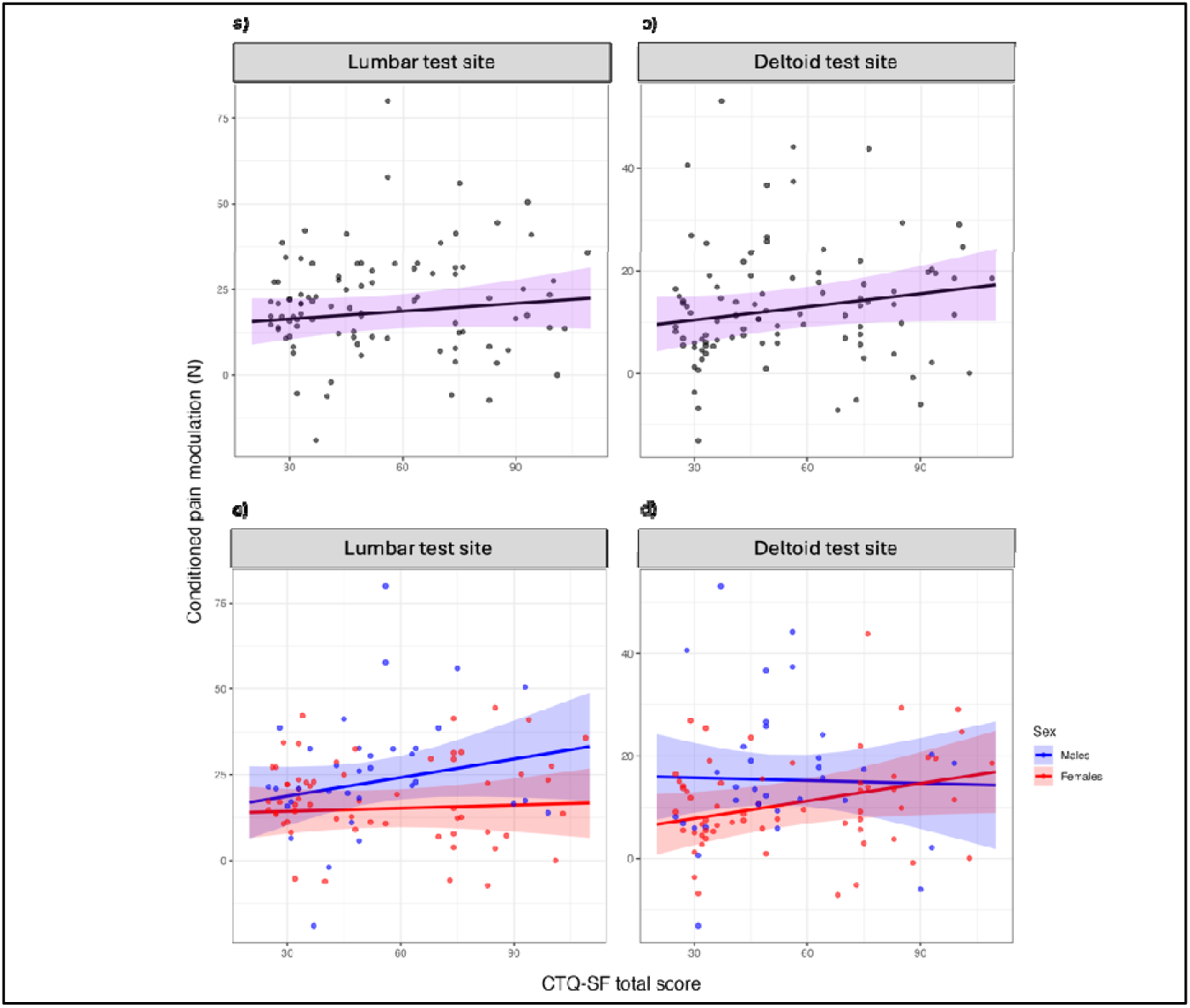
Plot of the association between the Childhood Trauma Questionnaire-Short Form (CTQ-SF) total score and conditioned pain modulation. Plots show the observed values (dots) and the predicted regression line and 95% confidence interval (ribbon) for the covariate-adjusted model for the full cohort (plots a and b) and with sex as an interaction term (plots c and d).

**Table 6:** Summary of the relationship between childhood adversity (CTQ-SF total score) and conditioned pain modulation.

| <u>Predictors</u> | Unadjusted model of main effect |  |  | Unadjusted model with sex interaction term |  |  | Adjusted model with sex interaction term |  |  |
| --- | --- | --- | --- | --- | --- | --- | --- | --- | --- |
|  | <u>Estimates</u> | <u>95% CI</u> | <u>p</u> | <u>Estimates</u> | <u>95% CI</u> | <u>p</u> | <u>Estimates</u> | <u>95% CI</u> | <u>p</u> |
| Intercept | 18.29 | 12.45; 24.13 | **<0.001** | 20.04 | 9.61; 30.47 | **<0.001** | 16.07 | 3.57; 28.57 | **0.01** |
| CTQ-SF score | 0.05 | -0.04; 0.15 | 0.29 | 0.10 | -0.08; 0.29 | 0.28 | 0.19 | -0.09; 0.28 | 0.31 |
| Site: deltoid vs lumbar | -8.07 | -11.04; -5.09 | **<0.001** | -8.07 | -11.04; -5.09 | **<0.001** | -8.07 | -11.04; -5.09 | **<0.001** |
| Sex: female vs male |  |  |  | -3.9 | -16.00; 8.4 | 0.53 | -5.39 | -17.48; 6.70 | 0.38 |
| CTQ-SF score x sex (female) |  |  |  | -0.05 | -0.27; 0.16 | 0.61 | -0.01 | -0.23; 0.20 | 0.90 |
| Age |  |  |  |  |  |  | 0.30 | -0.11; 0.39 | 0.27 |
| PSS score |  |  |  |  |  |  | -0.19 | -0.59; 0.20 | 0.34 |
| PSQI score |  |  |  |  |  |  | 0.20 | -0.47; 0.87 | 0.55 |
| Diagnosis of major depressive disorder |  |  |  |  |  |  | 6.47 | -1.90; 14.84 | 0.13 |
| Recent COVID-19 infection |  |  |  |  |  |  | 8.83 | 0.41; 17.24 | **0.04** |
| Recent acute illness |  |  |  |  |  |  | 2.38 | -2.06; 6.83 | 0.29 |
| Diagnosis of chronic illness |  |  |  |  |  |  | -6.72 | -14.17; 0.73 | 0.08 |
| P-values are rounded to two significant decimal points. |  |  |  |  |  |  |  |  |  |
| CTQ-SF = Childhood Trauma Questionnaire-short form; PSS = Perceived Stress Scale; PSQI = Pittsburgh Sleep Quality Index. |  |  |  |  |  |  |  |  |  |

### Benjamini–Hochberg correction for multiple tests

Although CTQ-SF total score was associated with the surface area of secondary hyperalgesia before correction for multiple comparisons (p = 0.043), this association did not remain statistically significant after adjustment (q = 0.17; Table 7). None of the remaining primary outcomes had a statistically significant relationship to childhood adversity before or after correction for multiple testing (Table 7).

**Table 7:** Benjamini–Hochberg correction for multiple tests

| Outcome | Estimate | 95%CI | p | q (BH) |
| --- | --- | --- | --- | --- |
| Surface area of secondary hyperalgesia | 0.32 | 0.01; 0.62 | **0.043** | 0.17 |
| Magnitude of secondary hyperalgesia | -0.10 | -0.28; 0.08 | 0.28 | 0.41 |
| Temporal summation |  |  | 0.73 | 0.73 |
| Conditioned pain modulation | 0.19 | -0.09; 0.28 | 0.31 | 0.41 |

### Exploratory analyses

The median [IQR] PSS score (possible range: 0 – 40) was 17 [12.5; 22.50] for the full cohort. There was no significant difference in the median [IQR] PSS score between males and females (males: 16.5 [12; 19]; females: 18 [14; 23]; *p* = 0.14; Table 2). Detailed explanations of the regression models and assessments of model assumptions are presented in Supplementary file: Section 4.1. There was no evidence of an association between PSS and the surface area or magnitude of secondary hyperalgesia, temporal summation, or CPM. There was also no evidence of an effect of sex, nor an effect of the interaction between PSS and sex.

## Discussion

This study examined the relationships of childhood adversity to four measures of experimental pain, and explored whether sex influenced these relationships in pain-free adults who had reported a range of severities of childhood adversity. More severe childhood adversity was associated with greater surface area of induced secondary hyperalgesia; however, this relationship did not remain after accounting for multiple testing. Childhood adversity was unrelated to the magnitude of induced secondary hyperalgesia, temporal summation, or CPM. No sex differences were observed. Recent stress was also unrelated to any of the four experimental pain outcomes.

Of the four experimental pain outcomes examined, the only evidence of a possible relationship with childhood adversity was for the surface area of induced secondary hyperalgesia, but this was neither robust nor sizeable. In the adjusted model, the effect size was equivalent to an average of 3.2 cm^2^ greater surface area for each 10-point difference in CTQ-SF score. Thus, our findings only tentatively suggest a subtle relationship between childhood adversity and the spatial extent of experimentally induced secondary hyperalgesia.

There is limited opportunity to compare this finding to existing literature because we could find no other published report on the relationship between childhood adversity and induced secondary hyperalgesia. Unpublished data from our lab, in males and females living with HIV with or without persistent pain, found no association between childhood adversity and induced secondary hyperalgesia. Some indirect support comes from one study in which a composite score incorporating both childhood adversity *and* recent stressful events was positively associated with the surface area of capsaicin-induced secondary hyperalgesia [70]—but the independent contribution of childhood adversity to that association cannot be determined. Taken together, the limited and mixed evidence indicates that the association observed in the present study requires further investigation.

The differing findings for the surface area and the magnitude of secondary hyperalgesia should not necessarily be interpreted as contradictory. Although these outcomes are commonly treated as comparable measures of experimentally induced secondary hyperalgesia, evidence suggests that they may not be equivalent. Meta-analytical evidence indicates that the magnitude and surface area of experimentally induced secondary hyperalgesia respond differently to pharmacological manipulations, suggesting that they may capture partly distinct physiological processes [4]. Other work by our group has also pointed out the lack of within-individual correlations between these two measures. While the physiological processes underlying the two measures are yet to be clarified, it seems that they may not entirely overlap, making it plausible that childhood adversity could be related to the spatial extent but not the magnitude of secondary hyperalgesia.

The lack of relationship between childhood adversity and temporal summation or CPM in this study aligns with many, although not all, of the relatively few and conflicting published studies on this topic. Of five studies including both males and females, three studies found that temporal summation was unrelated to childhood adversity [43, 58, 72], one found that childhood adversity was linked to greater temporal summation [71], and one found that childhood adversity was linked to *reduced* temporal summation [18]. Of two studies that investigated the relationship between childhood adversity and CPM, one found no relationship [32], whereas the other found that CPM was associated with childhood adversity but only in participants living in economically marginalised neighbourhoods [59]. Collectively, these inconsistent findings raise the possibility that contextual or individual factors may be necessary to reveal the physiological consequences of childhood adversity.

It seems possible that subsequent exposures, or other moderating factors, may influence whether a vulnerability arising from exposure to childhood adversity is expressed as altered pain modulation in adulthood. One potentially important moderator may be social support, which has been associated with reduced clinical and experimentally induced pain [31]. For example, participants receiving social support from a romantic partner or stranger during induction of secondary hyperalgesia showed a smaller surface area and magnitude of secondary hyperalgesia than those in a control condition [28, 29]. Social support may also buffer inflammatory processes that are associated with childhood adversity. Higher social support has been associated with lower circulating concentrations of pro-inflammatory cytokines [60], and one study found that social support moderated the relationship between childhood adversity and C-reactive protein [52]. That the relationship between childhood adversity and pain modulation may be modified by protective social factors calls for future work to consider whether socio-environmental factors act to influence the relationship between childhood adversity and pain.

Our findings add to, rather than resolve, the conflicting evidence about how childhood adversity relates to clinical persistent pain vs experimental pain sensitivity. While childhood adversity is consistently associated with an increased risk and greater severity of clinical persistent pain [10, 14, 15, 21, 22, 54, 69], the small existing body of experimental literature reports inconsistent findings across experimental outcomes of secondary hyperalgesia, temporal summation, and CPM. Although these outcomes probe selected components of nociceptive processing and are commonly interpreted as markers of vulnerability to persistent pain, their relationship to clinically meaningful pain outcomes remains tenuous [17]. It may therefore be more productive for future research to focus on non-nociceptive, environmental, and/or social mechanisms, as well as their interactions with each other and with the nociceptive system, as potential pathways from childhood adversity to increased risk of clinical persistent pain.

### Strengths and limitations

This study provides deeper insight to the relationship between childhood adversity and pain modulation than we have seen before [43, 70–72], specifically because we assessed four measures of pain modulation, including processes dominated by facilitation and inhibition, and two that likely reflect both. This allowed us to compare relationships across measures. We were also able to explore the relationship of recent stress to these four measures.

There are three key limitations of this study. First, the distribution of males to females in this cohort was unequal (n = 34 male; n = 61 female), which broadly reflects the real-world prevalence of persistent pain but will have increased uncertainty in the effect estimates for the sex comparisons. Furthermore, although biological sex was included in our analyses, we did not collect data on hormonal status or examine whether hormonal factors influenced our outcomes as recommended by the PAINDIFF group [19]. Second, although the CTQ-SF is one of the most widely used and validated tools globally [23], including in South Africa [12, 30, 56, 57], it is an event-based measure that fails to capture the subjective magnitude or longevity of the impact of the adverse experience. Third, our assessor was unblinded to the research hypotheses, and this secondary analysis did not have a prespecified locked protocol. To mitigate this, we have provided the analysis code and de-identified data at https://osf.io/y34pa/.

## Conclusion

In this sample of healthy adults with a range of self-reported severity of childhood adversity, more severe childhood adversity was associated with a greater surface area of induced secondary hyperalgesia; however, this association was not upheld when accounting for multiple testing. Childhood adversity was also unrelated to the magnitude of induced secondary hyperalgesia, temporal summation, or CPM. Recent stress was also unrelated to any of the four experimental pain outcomes, and there were no differences between sexes.

## Data Availability

The analysis code and de-identified data are at https://osf.io/y34pa/.

## Acknowledgements

The authors thank Justin Pead (University of Cape Town) for his technical assistance with automating the high-frequency electrical stimulation trains, Dr Kessie Govender for making the cathodes, and Mathijs Franssen (KU Leuven) for research support.

## Funding details

GJB was supported by postgraduate scholarships from African Pain Research Initiative (University of Cape Town), PainSA, the National Research Foundation (South Africa), the Oppenheimer Memorial Trust, and a Postgraduate Publication Incentive Award (University of Cape Town).

LM is supported by a postgraduate scholarship from the University of Cape Town and the National Research Foundation (South Africa).

MRH is supported by an Australian Research Council Future Fellowship (FT180100565). RP is supported by the National Research Foundation of South Africa as a rated researcher. VJM was supported by the Fogarty International Center of the National Institutes of Health (award K43TW011442).

## Conflicts of interest

GJB occasionally receives speakers’ fees for talks on pain and rehabilitation from the South African National Department of Health, PainSA, and the South African Society of Physiotherapy.

LM has no conflicts of interest related to this work.

MRH is a director of the not-for-profit organisation Australian Pain Research Solution Alliance, Chair of the Safeguarding Australia through Biotechnology Response and Engagement (SABRE) Alliance, a member of the Prime Minister’s National Science and Technology Council, and a Director of the Australia’s Economic Accelerator Advisory Board. RP receives speakers’ fees for talks on pain and rehabilitation from the not-for-profit organisation Train Pain Academy and the Haleon and Faircape Group, is a director of Train

Pain Academy, and serves as a councillor for the International Association for the Study of Pain.

VJM is an associate director of, and occasionally receives speakers’ fees from, the not-for-profit organisation, Train Pain Academy.

## Authorship contributions

GJB, VJM, MRH, and RP conceived and initiated the study. GJB led the execution of the study. LM contributed to data collection. VJM contributed to data analysis. All authors contributed to data interpretation and critically revised the manuscript.

## Notes

### Clinical Protocols

https://osf.io/y34pa/

### Author Declarations

Ethical approval was obtain from the University of Cape Town, Faculty of Health Sciences Human Research Ethics Committee (reference number: 560/2021)

### Summary of Updates

Following constructive peer review, we revised the statistical analysis to account for multiple testing across our four primary hypotheses. Specifically, we applied a multiple-comparison correction to the primary analyses and revised the Results accordingly. We also updated the Discussion to reflect the corrected results and their implications for the interpretation and conclusions of the study. These revisions improve the statistical rigour of the analysis and ensure that the interpretation appropriately accounts for the number of primary hypotheses tested.

