## Supplementary file for "How does adversity in childhood relate to pain in adulthood? Findings across four experimental tests of pain modulation"

### 1. STROBE checklist

Table S1: STROBE checklist.

|  | Item No | Recommendation | Page  No |
| --- | --- | --- | --- |
| **Title and abstract** | 1 | (*a*) Indicate the study’s design with a commonly used term in the title or the abstract | 2 |
|  |  | (*b*) Provide in the abstract an informative and balanced summary of what was done and what was found | 2 |
| Introduction | | | |
| Background/rationale | 2 | Explain the scientific background and rationale for the investigation being reported | 3 & 4 |
| Objectives | 3 | State specific objectives, including any prespecified hypotheses | 4 |
| Methods | | | |
| Study design | 4 | Present key elements of study design early in the paper | 4 & 5 |
| Setting | 5 | Describe the setting, locations, and relevant dates, including periods of recruitment, exposure, follow-up, and data collection | 5 – 9 |
| Participants | 6 | (*a*) *Cohort study*—Give the eligibility criteria, and the sources and methods of selection of participants. Describe methods of follow-up  *Case-control study*—Give the eligibility criteria, and the sources and methods of case ascertainment and control selection. Give the rationale for the choice of cases and controls  *Cross-sectional study*—Give the eligibility criteria, and the sources and methods of selection of participants | 5 (Table 1) |
|  |  | (*b*) *Cohort study*—For matched studies, give matching criteria and number of exposed and unexposed  *Case-control study*—For matched studies, give matching criteria and the number of controls per case | N/A |
| Variables | 7 | Clearly define all outcomes, exposures, predictors, potential confounders, and effect modifiers. Give diagnostic criteria, if applicable | 7 & 8 |
| Data sources/ measurement | 8* | For each variable of interest, give sources of data and details of methods of assessment (measurement). Describe comparability of assessment methods if there is more than one group | 7 & 8 |
| Bias | 9 | Describe any efforts to address potential sources of bias | 20 |
| Study size | 10 | Explain how the study size was arrived at | 8 |
| Quantitative variables | 11 | Explain how quantitative variables were handled in the analyses. If applicable, describe which groupings were chosen and why | 8 |
| Statistical methods | 12 | (*a*) Describe all statistical methods, including those used to control for confounding | 8 |
|  |  | (*b*) Describe any methods used to examine subgroups and interactions | 8 & 9 |
|  |  | (*c*) Explain how missing data were addressed | 12 |
|  |  | (*d*) *Cohort study*—If applicable, explain how loss to follow-up was addressed  *Case-control study*—If applicable, explain how matching of cases and controls was addressed  *Cross-sectional study*—If applicable, describe analytical methods taking account of sampling strategy |  |
|  |  | (*e*) Describe any sensitivity analyses | 10 |
| **Results** |  |  |  |
| Participants | 13* | (a) Report numbers of individuals at each stage of study—eg numbers potentially eligible, examined for eligibility, confirmed eligible, included in the study, completing follow-up, and analysed | 11 |
|  |  | (b) Give reasons for non-participation at each stage | 11 |
|  |  | (c) Consider use of a flow diagram | N/A |
| Descriptive data | 14* | (a) Give characteristics of study participants (eg demographic, clinical, social) and information on exposures and potential confounders | 11 (Table 2) |
|  |  | (b) Indicate number of participants with missing data for each variable of interest | 11 |
|  |  | (c) *Cohort study*—Summarise follow-up time (eg, average and total amount) | N/A |
| Outcome data | 15* | *Cohort study*—Report numbers of outcome events or summary measures over time | N/A |
|  |  | *Case-control study—*Report numbers in each exposure category, or summary measures of exposure | N/A |
|  |  | *Cross-sectional study—*Report numbers of outcome events or summary measures | 11 (Table 2) |
| Main results | 16 | (*a*) Give unadjusted estimates and, if applicable, confounder-adjusted estimates and their precision (eg, 95% confidence interval). Make clear which confounders were adjusted for and why they were included | 12 – 15 |
|  |  | (*b*) Report category boundaries when continuous variables were categorized | N/A |
|  |  | (*c*) If relevant, consider translating estimates of relative risk into absolute risk for a meaningful time period | N/A |
| Other analysises | 17 | Report other analyses done—eg analyses of subgroups and interactions, and sensitivity analyses | 15 and Supplementary file |
| **Discussion** |  |  |  |
| Key results | 18 | Summarise key results with reference to study objectives | 16 |
| Limitations | 19 | Discuss limitations of the study, taking into account sources of potential bias or imprecision. Discuss both direction and magnitude of any potential bias | 19 & 20 |
| Interpretation | 20 | Give a cautious overall interpretation of results considering objectives, limitations, multiplicity of analyses, results from similar studies, and other relevant evidence | 16 – 19 |
| Generalisability | 21 | Discuss the generalisability (external validity) of the study results | 20 |
| **Other information** |  |  |  |
| Funding | 22 | Give the source of funding and the role of the funders for the present study and, if applicable, for the original study on which the present article is based | 21 |

#### Outcome measures for potential confounders

Positive childhood experiences

Participants reported on positive childhood experiences using the Positive Childhood Experiences Questionnaire [57]. This questionnaire has seven items to which participants endorsed answers using a 5-point Likert scale (0 – never; 1 – rarely; 2 – sometimes; 3 – often; 4 – very often). The total score was computed as the sum of all seven item scores. Positive childhood experiences may buffer the effects of adverse childhood experiences [57]. We tested whether score on the positive childhood experiences questionnaire influenced the relationship childhood adversity and inflammatory reactivity.

Long-term stress

Participants completed the 10-item Perceived Stress Scale [58] by rating each item (e.g. *In the last month, how often have you been able to control the irritations in your life)* on a 5-point Likert scale (1 – never; 2 – almost never; 3 – sometimes; 4 – fairly often; 5 – very often). The total score was computed as the sum of all 10 item scores. Long-term stress alters inflammatory reactivity, as seen by high levels of IL-6 in people exposed to chronic stress [59-65]. Further, individuals exposed to long-term stress of caring for a spouse with dementia showed lower levels of IL-2 after influenza vaccine immune provocation than age- and sex-matched controls [66]. Therefore, we tested whether long-term stress was a potential confounder of inflammatory reactivity, surface area and magnitude of secondary hypersensitivity, CPM, and/or TS.

Depression and anxiety

We screened for a lifetime history of diagnosed major depressive disorder, as well as screened for current symptoms of depression and anxiety using the well-validated and reliable Patient Health Questionnaire-4 [67]. This questionnaire has 4-items to which answers were endorsed using a 4-point Likert scale (0 – not at all; 1 – several days; 2 – more than half the days; 3 – nearly every day). The sum of the first two items screen for anxiety and the sum of the last two items screen for depression (>3 is considered positive for either of these subscales). Depression alters inflammatory reactivity, as seen by a meta-analysis of data from 3212 people reporting higher levels of IL-6 and TNF-α, as well as other cytokines, in peripheral blood of people with major depressive disorder, than in healthy controls [68]. Anxiety is also associated with higher levels of IL-6 and TNF-α [69]. Therefore, we tested whether depression and/or anxiety (total screening score on Patient Health Questionnaire-4, and previous diagnosis of major depressive disorder) were potential confounders of inflammatory reactivity, surface area and magnitude of secondary hypersensitivity, CPM, and/or TS.

Asthma

Participants self-reported history of diagnosed asthma. Asthma alters inflammatory reactivity, as seen by preliminary, unpublished data from our research team. Further, asthma history is associated with reduced levels of the largely anti-inflammatory cytokine IL-10 [70] in LPS-stimulated blood culture, and asthma severity is negatively associated with levels of IL-10 [71]. Therefore, we tested whether diagnosed asthma was a potential confounder of inflammatory reactivity.

COVID-19 infection

Participants self-reported history of COVID-19 infection in the six months preceding participation. In participants with a history of COVID-19 infection, we collected data on the timing and severity of known COVID-19 infection and severity of long-COVID symptoms. Recent COVID-19 infection may alter inflammatory reactivity, and could confound interpretation of IL-6 and TNF-α levels in stimulated and unstimulated blood samples. COVID-19 infection stimulates a cytokine-driven inflammatory response that can build into a ‘cytokine storm’ with profound and lasting effects [72, 73]. Further, persistent symptoms after acute COVID-19 infection, termed “long-COVID” are associated with pro-inflammatory cytokine activation [74, 75]. Therefore, history of COVID-19 infection may be associated with inflammatory reactivity. Unfortunately, we were unable to account for undiagnosed COVID-19 infection, which is suspected to be common although the inflammatory consequences are unknown. We tested whether COVID-19 infection was a potential confounder of inflammatory reactivity.

Chronic and recent acute illnesses

Participants self-reported chronic illnesses and any recent acute infections in the six months preceding participation, and we tested whether chronic illnesses (e.g. HIV) and recent acute illnesses (e.g. influenza) was a potential confounder of inflammatory reactivity.

Sleep

Participants reported sleep quality using the Pittsburgh Sleep Quality Index [76], which consists of 19 self-rated items, the sum of which provides a total score out of 21. Sleep deprivation and/or disturbance can alter inflammatory and neural reactivity, as shown by data from humans under resting and challenged states. A large meta-analysis of data from 3000 participants reported a strong positive association between recent sleep disturbance and circulating IL-6 levels [77]. Additionally, poor sleep quality was associated with higher levels of IL-6 after *in vivo* stimulation using LPS in African American women [78]. Further, one night of sleep deprivation increased the area of experimentally induced secondary hyperalgesia in healthy male, but not female, subjects [79]. Therefore, we tested whether sleep deprivation (total score out of 21) was a potential confounder of inflammatory reactivity, surface area and magnitude of secondary hypersensitivity, CPM, and/or TS.

#### 3. Plots of model assumptions

##### a. Outcome: surface area of secondary hyperalgesia

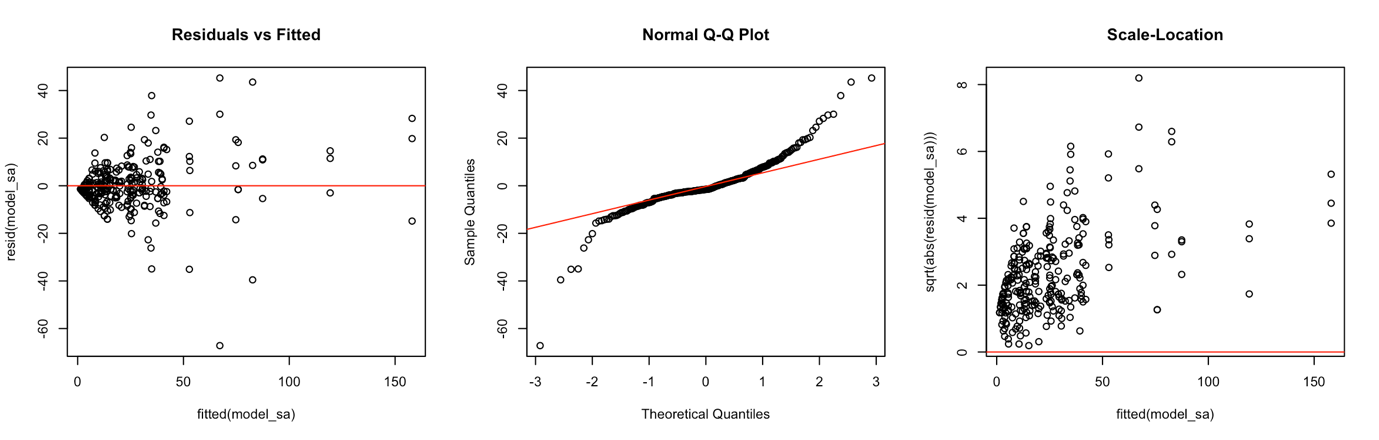

Figure S1: Plots of model assumptions for the conventional mixed-effects linear regression investigating the relationship between Childhood Trauma Questionnaire-Short Form (CTQ-SF) and surface area of secondary hyperalgesia.

##### b. Outcome: magnitude of secondary hyperalgesia

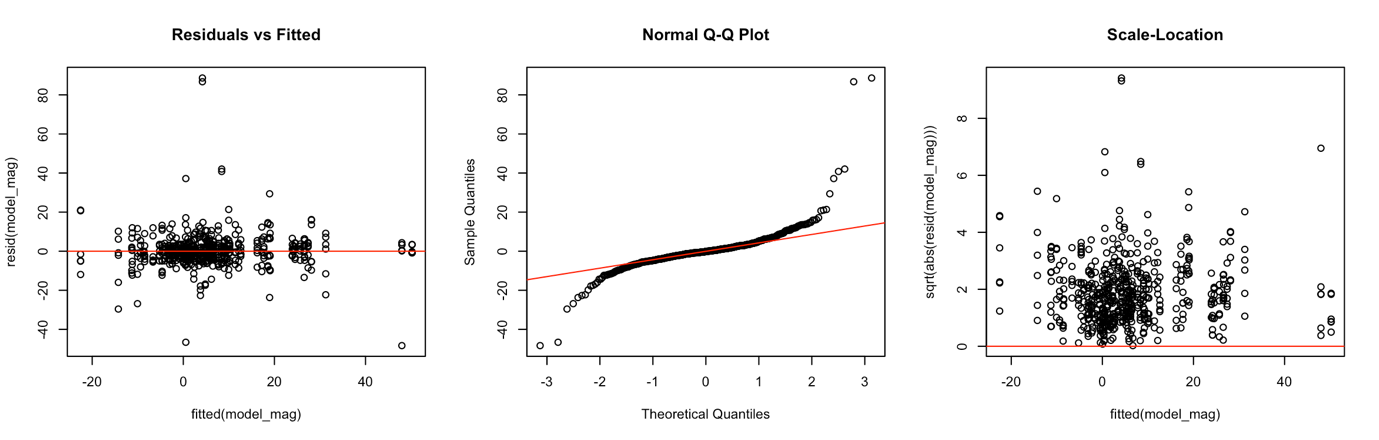

Figure S2: Plots of model assumptions for the conventional mixed-effects linear regression investigating the relationship between Childhood Trauma Questionnaire-Short Form (CTQ-SF) and magnitude of secondary hyperalgesia.

##### c. Outcome: temporal summation

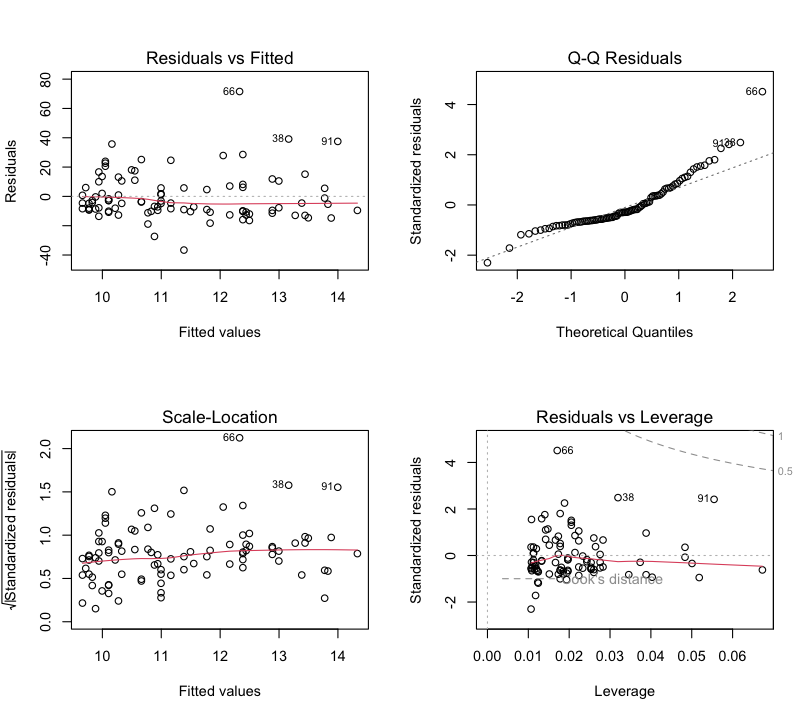

Figure S3: Plots of model assumptions for the conventional linear regression investigating the relationship between Childhood Trauma Questionnaire-Short Form (CTQ-SF) and temporal summation at the lumbar site.

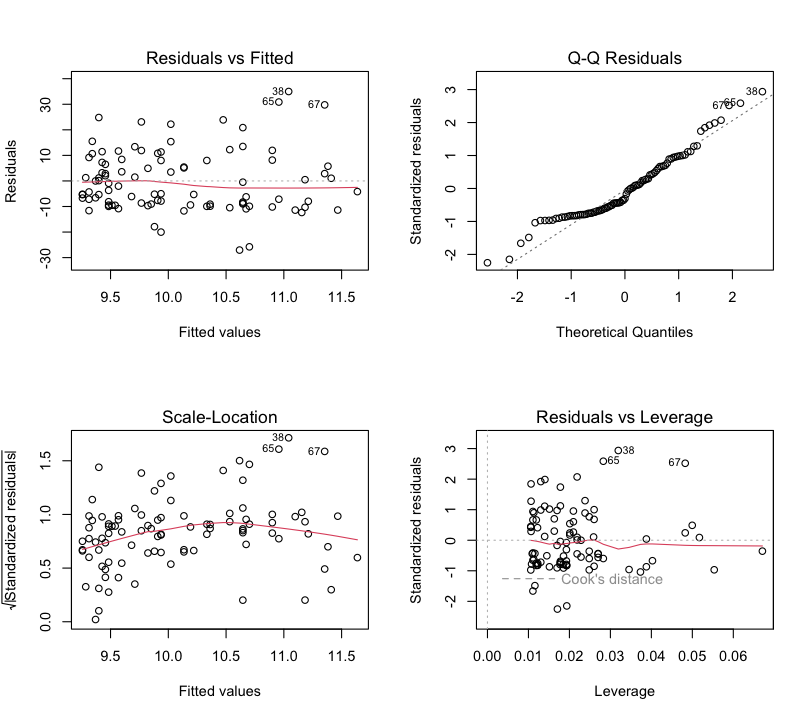

Figure S4: Plots of model assumptions for the conventional linear regression investigating the relationship between Childhood Trauma Questionnaire-Short Form (CTQ-SF) and temporal summation at the deltoid site.

##### d. Outcome: conditioned pain modulation

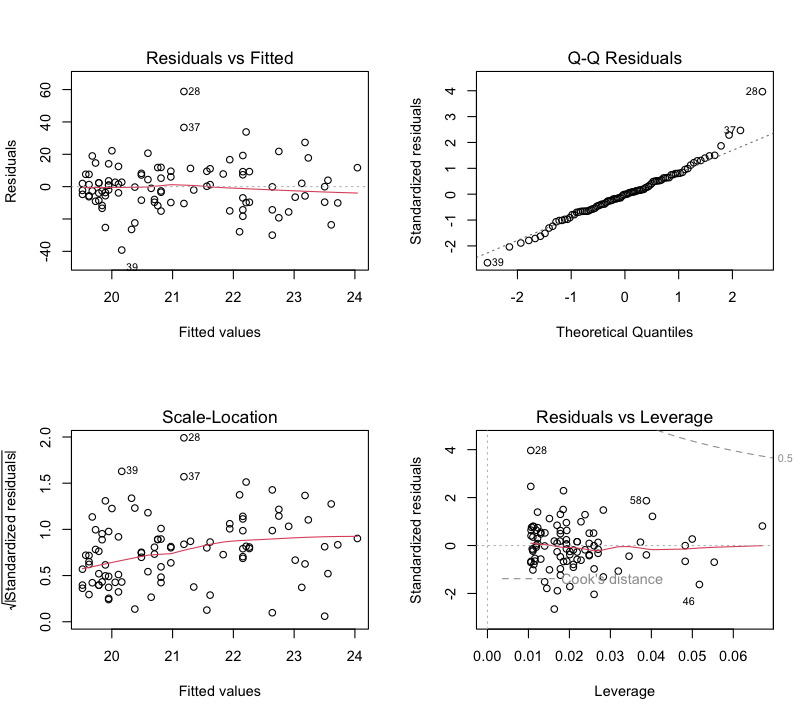

Figure S5: Plots of model assumptions for the conventional linear regression investigating the relationship between Childhood Trauma Questionnaire-Short Form (CTQ-SF) and conditioned pain modulation at the lumbar site.

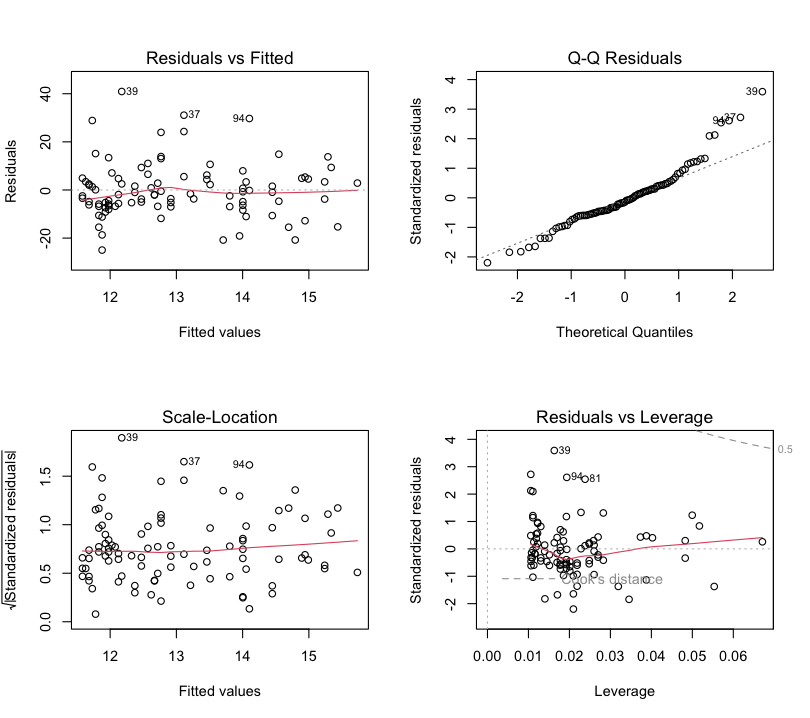

Figure S6: Plots of model assumptions for the conventional linear regression investigating the relationship between Childhood Trauma Questionnaire-Short Form (CTQ-SF) and conditioned pain modulation at the deltoid site.

#### 4. Exploratory analyses

##### Predictor: Perceived Stress Scale (PSS)

###### a. Outcome: surface area of secondary hyperalgesia

The unadjusted model found no evidence of an association between the PSS and the surface area of secondary hyperalgesia (β= 0.05 [95%CI: -0.46; 0.57], *p* = 0.83). When sex was included as an interaction term, both the unadjusted (β= 0.30 [95%CI: -0.63; 1.23], *p* = 0.53) and the covariate-adjusted models (β= 0.14 [95%CI: -0.84; 1.12], *p* = 0.78) found no evidence of an association between PSS and the surface area of secondary hyperalgesia, and no evidence of an effect of sex, nor an interaction between PSS and sex (Fig S8 and Table S2).

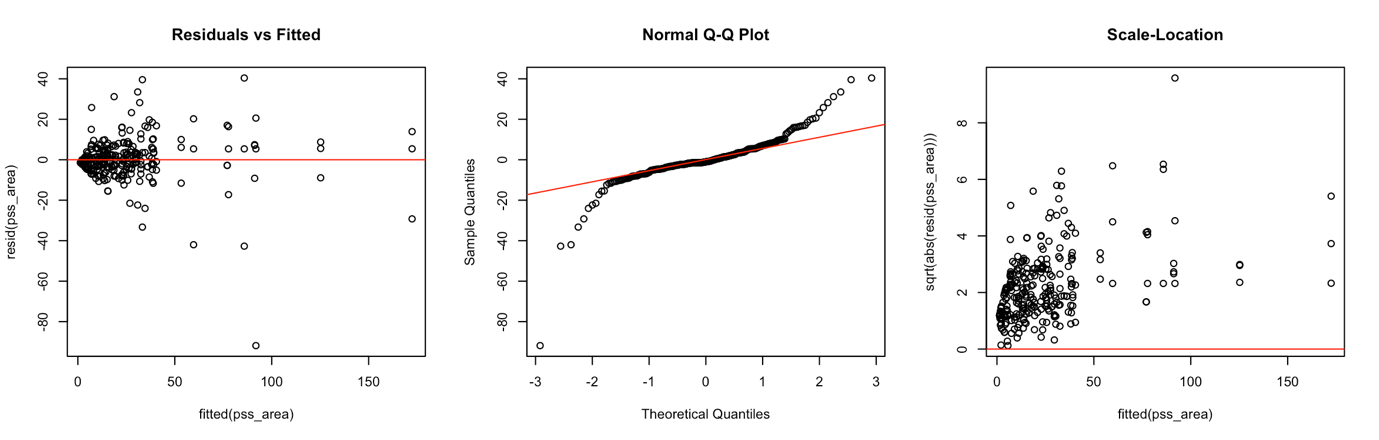

Figure S7: Plots of model assumptions for the conventional mixed-effects linear regression investigating the relationship between the Perceived Stress Scale (PSS) and the surface area of secondary hyperalgesia.

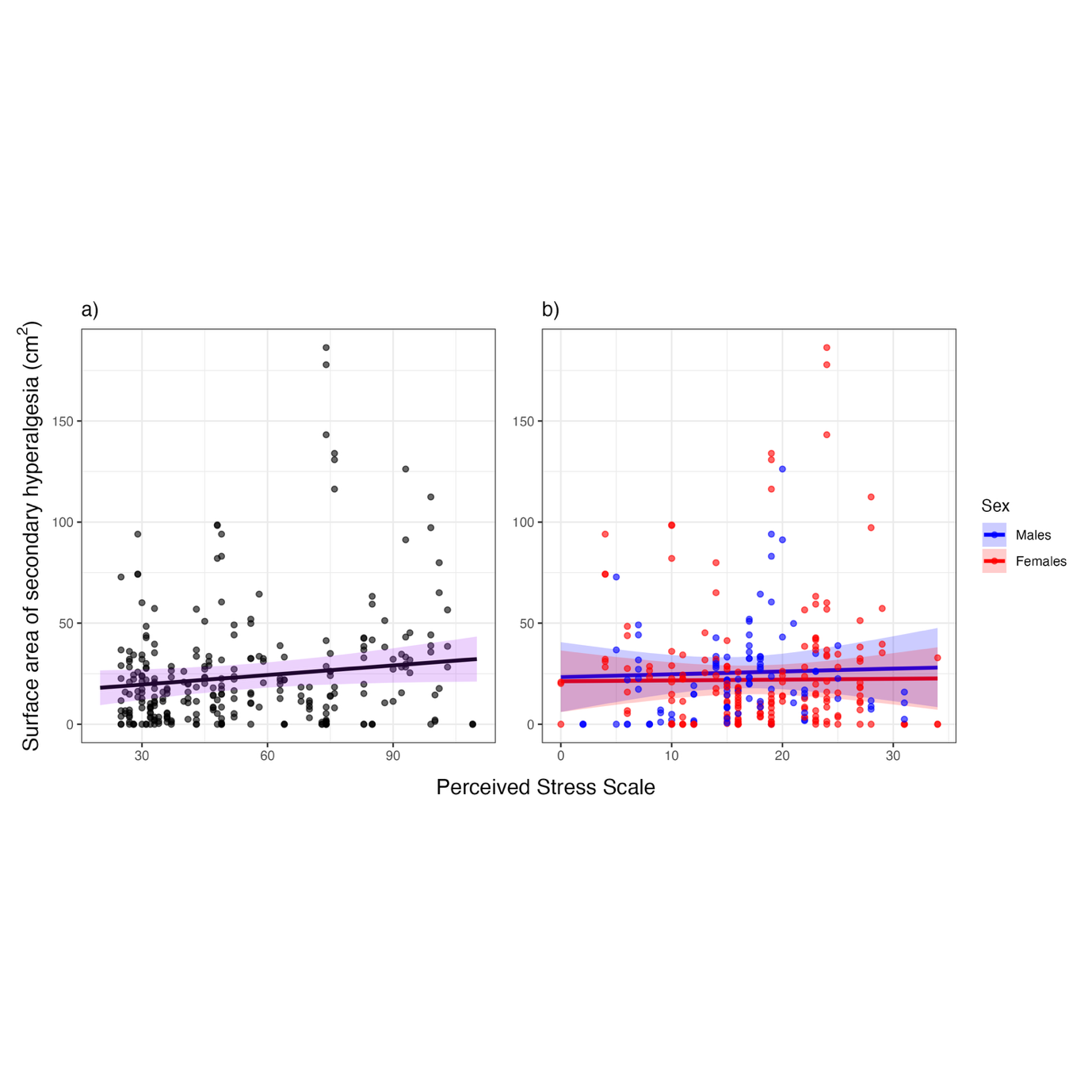

Figure S8: Plot of the association between the Perceived Stress Scale (PSS) and the surface area of secondary hyperalgesia. Plots show the observed values (dots) and the predicted regression line and 95% confidence interval (ribbon) for the covariate-adjusted model for the full cohort (plot a) and with sex as an interaction term (plot b).

Table S2: Summary of the relationship between recent stress (Perceived stress Scale: PSS) and surface area of induced secondary hyperalgesia.

| Predictors | **Unadjusted model of main effect** | | | **Unadjusted model with sex interaction term** | | | **Adjusted model with sex interaction term** | | |
| --- | --- | --- | --- | --- | --- | --- | --- | --- | --- |
|  | Estimates | 95% CI | *p* | Estimates | 95% CI | *p* | Estimates | 95% CI | *p* |
| Intercept | 18.54 | 9.09; 27.99 | **<0.001** | 15.69 | -0.10; 31.49 | 0.052 | 13.42 | -12.35; 39.18 | 0.31 |
| PSS score | 0.05 | -0.46; 0.57 | 0.83 | 0.30 | -0.63; 1.23 | 0.53 | 0.14 | -0.84; 1.12 | 0.78 |
| Sex (female) |  |  |  | 4.13 | -15.77; 24.04 | 0.68 | -2.08 | -22.86; 18.71 | 0.84 |
| PSS score x sex (female) |  |  |  | -0.34 | -1.46; 0.78 | 0.55 | -0.10 | -1.24; 1.04 | 0.87 |
| Age |  |  |  |  |  |  | 0.47 | 0.05; 0.90 | **0.030** |
| Mean SPARS rating of HFS induction |  |  |  |  |  |  | -0.08 | -0.41; 0.26 | 0.65 |
| HFS current intensity |  |  |  |  |  |  | -36.71 | -81.24; 7.81 | 0.11 |
| CTQ-SF total score |  |  |  |  |  |  | 0.14 | -0.03; 0.32 | 0.10 |
| PSQI score |  |  |  |  |  |  | -0.20 | -1.36; 0.96 | 0.73 |
| Diagnosis of major depressive disorder |  |  |  |  |  |  | 0.60 | -13.67; 14.88 | 0.93 |
| Recent COVID-19 infection |  |  |  |  |  |  | 6.29 | -9.22; 21.80 | 0.43 |
| Recent acute illness |  |  |  |  |  |  | -4.89 | -13.24; 3.45 | 0.25 |
| Diagnosis of chronic illness |  |  |  |  |  |  | -7.37 | -19.98; 5.24 | 0.25 |
| **Random effects** | | | | | | | | | |
| σ^2^ | 66.30 | | | 66.06 | | | 65.37 | | |
| τ_00_ | 274.50_study_id_ | | | 278.37_study_id_ | | | 269.98_study_id_ | | |
| ICC | 0.81 | | | 0.81 | | | 0.81 | | |
| N | 95_study_id_ | | | 95_study_id_ | | | 95_study_id_ | | |
| Observations | 285 | | | 285 | | | 285 | | |
| Marginal R^2^ / Conditional R^2^ | 0.000/0.806 | | | 0.005/0.809 | | | 0.132/0.831 | | |
| PSS = Perceived Stress Scale; CTQ-SF = Childhood Trauma Questionnaire-short form; HFS = high-frequency electrical stimulation; PSQI = Pittsburgh Sleep Quality Index. | | | | | | | | | |

###### b. Outcome: magnitude of secondary hyperalgesia

The unadjusted model found no evidence of an association between PSS and the magnitude of SH (β= 0.05 [95%CI: -0.25; 0.35], *p* = 0.75). When sex was included as an interaction term, both the unadjusted (β= -0.02 [95%CI: -0.57; 0.54], *p* = 0.95) and the covariate-adjusted models (β= 0.30 [95%CI: -0.28; 0.88], *p* = 0.32) found no evidence of an association between PSS and the magnitude of SH, and no evidence of an effect of sex, nor an interaction between PSS and sex (Fig S10 and Table S3).

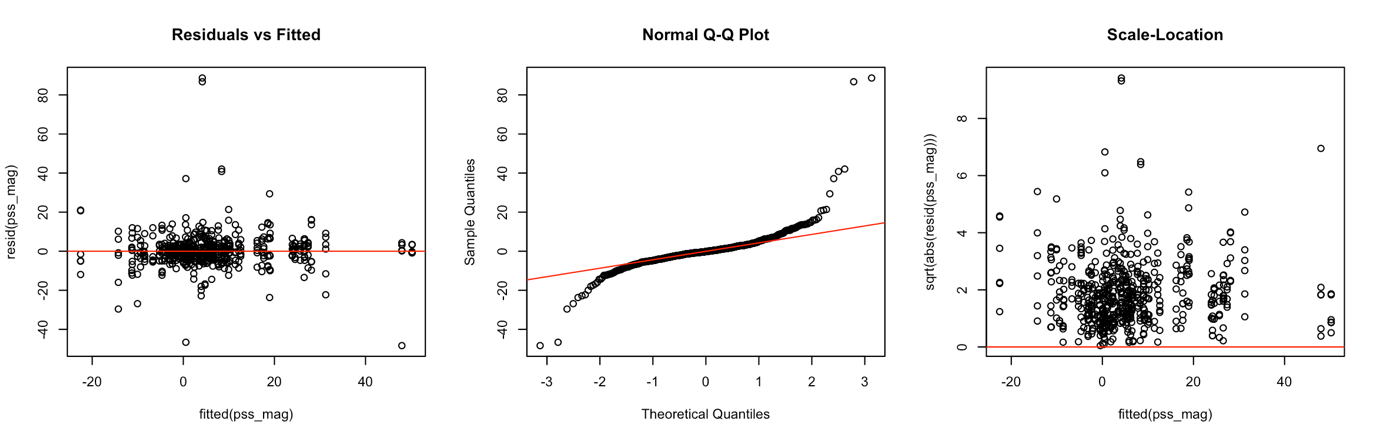

Figure S9: Plots of model assumptions for the conventional mixed-effects linear regression investigating the relationship between the Perceived Stress Scale (PSS) and the magnitude of secondary hyperalgesia.

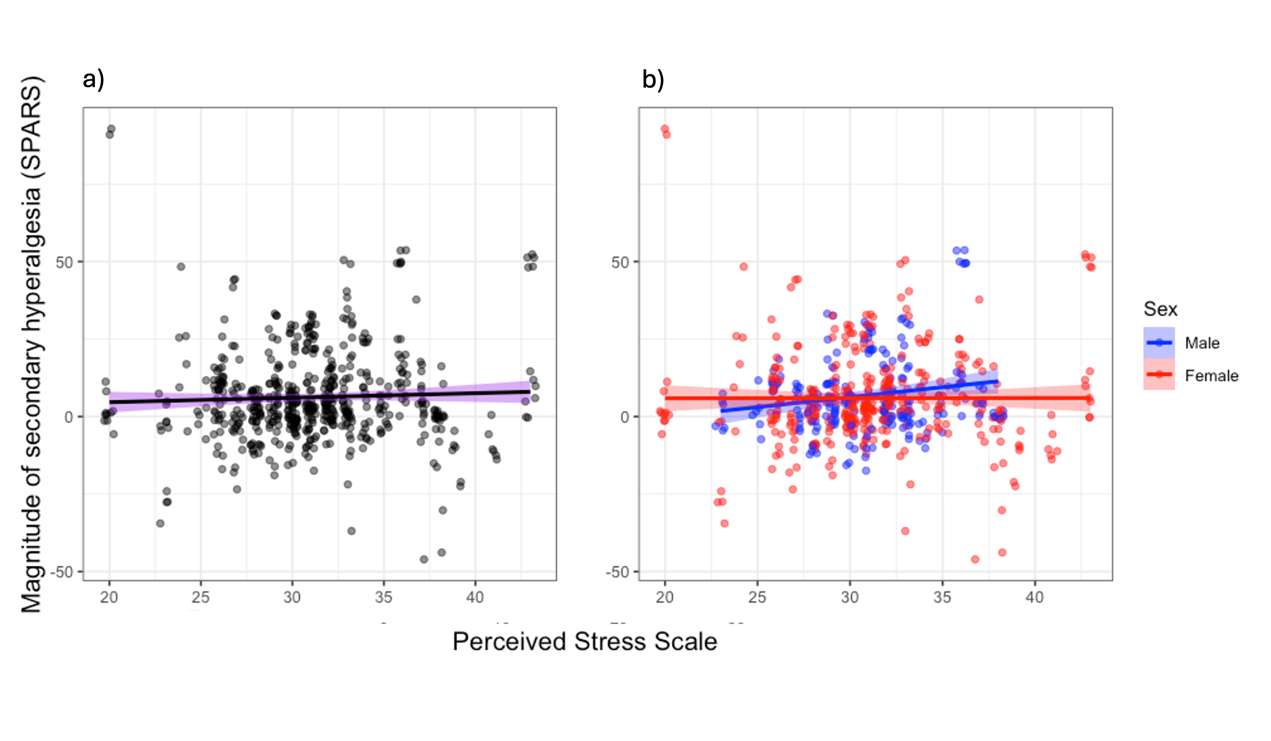

Figure S10: Plot of the association between the Perceived Stress Scale (PSS) and the magnitude of secondary hyperalgesia. Plots show the observed values (dots) and the predicted regression line and 95% confidence interval (ribbon) for the covariate-adjusted model for the full cohort (plot a) and with sex as an interaction term (plot b). Y axes have been truncated but range from -100 to +100.

Table S3: Summary of the relationship between recent stress (Perceived stress Scale: PSS) and magnitude of induced secondary hyperalgesia.

| Predictors | **Unadjusted model of main effect** | | | **Unadjusted model with sex interaction term** | | | **Adjusted model with sex interaction term** | | |
| --- | --- | --- | --- | --- | --- | --- | --- | --- | --- |
|  | Estimates | 95% CI | *p* | Estimates | 95% CI | *p* | Estimates | 95% CI | *p* |
| Intercept | 3.86 | -1.66; 9.39 | 0.17 | 5.40 | -3.98; 14.78 | 0.26 | 5.74 | -9.48; 20.95 | 0.46 |
| PSS score | 0.05 | -0.25; 0.35 | 0.75 | -0.02 | -0.57; 0.54 | 0.95 | 0.30 | -0.28; 0.88 | 0.32 |
| Sex (female) |  |  |  | -2.47 | -14.28; 9.35 | 0.68 | -5.26 | -17.53; 7.01 | 0.40 |
| PSS score x sex (female) |  |  |  | 0.10 | -0.56; 0.77 | 0.76 | 0.26 | -0.42; 0.93 | 0.45 |
| Age |  |  |  |  |  |  | 0.12 | -0.13; 0.37 | 0.35 |
| Mean SPARS rating of HFS induction |  |  |  |  |  |  | -0.17 | -0.37; 0.03 | 0.091 |
| HFS current intensity |  |  |  |  |  |  | 17.91 | -8.38; 44.20 | 0.18 |
| CTQ-SF total score |  |  |  |  |  |  | -0.03 | -0.13; 0.07 | 0.61 |
| PSQI score |  |  |  |  |  |  | -0.43 | -1.11; 0.26 | 0.22 |
| Diagnosis of major depressive disorder |  |  |  |  |  |  | 5.14 | -3.28; 13.57 | 0.23 |
| Recent COVID-19 infection |  |  |  |  |  |  | 4.61 | -4.55; 13.77 | 0.32 |
| Recent acute illness |  |  |  |  |  |  | -5.45 | -10.38; -0.53 | **0.030** |
| Diagnosis of chronic illness |  |  |  |  |  |  | 4.76 | -2.68; 12.21 | 0.21 |
| **Random effects** | | | | | | | | | |
| σ^2^ | 30.38 | | | 30.37 | | | 30.45 | | |
| τ_00_ | 0.00_modality:study_id_ | | | 0.00_modality:study_id_ | | | 0.00_modality:study_id_ | | |
|  | 96.79_study_id_ | | | 101.34_study_id_ | | | 97.10_study_id_ | | |
| N | 2_modality_ | | | 2_modality_ | | | 2_modality_ | | |
|  | 95_study_id_ | | | 95_study_id_ | | | 95_study_id_ | | |
| Observations | 570 | | | 570 | | | 570 | | |
| Marginal R^2^ / Conditional R^2^ | 0.004/NA | | | 0.011/NA | | | 0.464/NA | | |
| PSS = Perceived Stress Scale; CTQ-SF = Childhood Trauma Questionnaire-short form; HFS = high-frequency electrical stimulation; PSQI = Pittsburgh Sleep Quality Index. | | | | | | | | | |

###### c. Outcome: temporal summation

The unadjusted model satisfied the underlying assumptions of linear regression (Figs S11 and S12) and found no evidence of an association between the PSS and TS at either the lumbar (β=0.23 [95%CI: -0.65; 1.11], *p* = 0.60) or the deltoid site (β=0.19 [95%CI: -0.29; 0.88], *p* = 0.59). When sex was included as an interaction term, both the unadjusted and the covariate-adjusted models found no evidence of an association between PSS and TS at either the lumber or the deltoid site, and there was no evidence of an effect of sex, or an effect of the interaction between PSS and sex (Fig S13 and Tables S4 and S5).

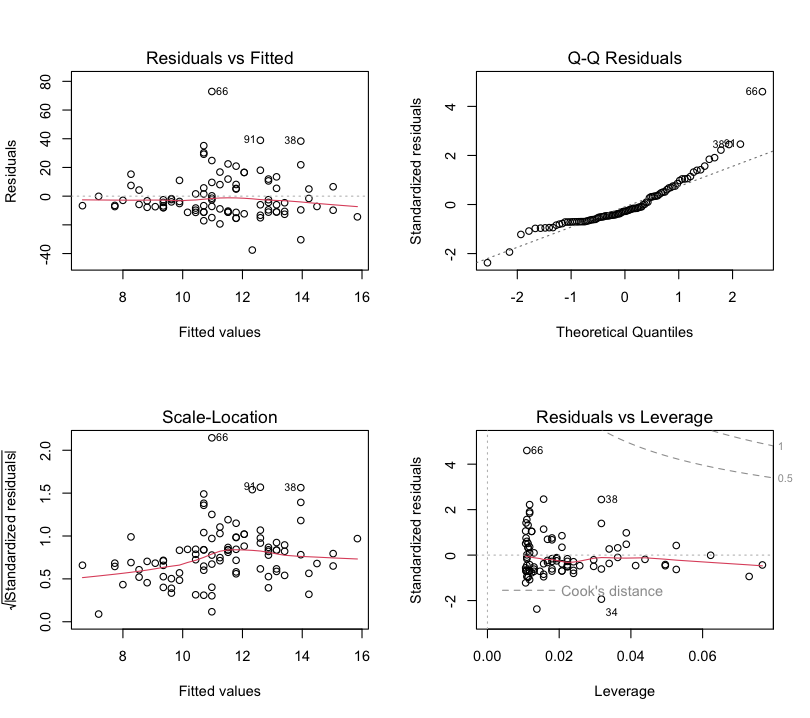

Figure S11: Plots of model assumptions for the conventional linear regression investigating the relationship between Perceived Stress Scale (PSS) and temporal summation at the lumbar site.

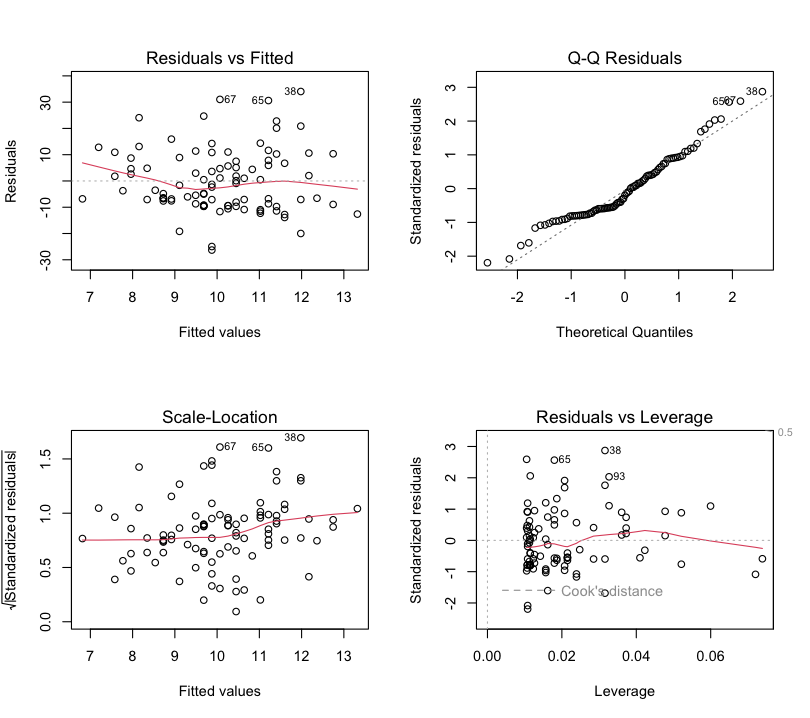

Figure S12: Plots of model assumptions for the conventional linear regression investigating the relationship between Perceived Stress Scale (PSS) and temporal summation at the deltoid site

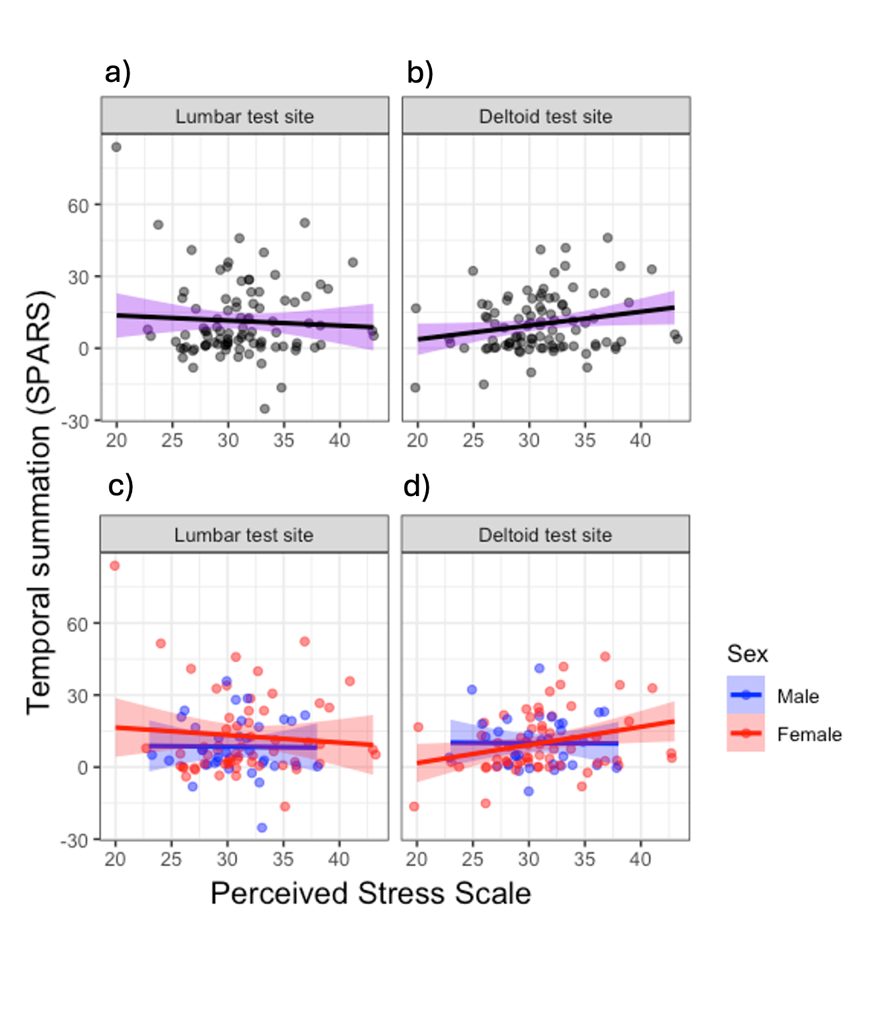

Figure S13: Plot of the association between the Perceived Stress Scale (PSS) and temporal summation. Plots show the observed values (dots) and the predicted regression line and 95% confidence interval (ribbon) for the covariate-adjusted model for the full cohort (plots a and b) and with sex as an interaction term (plots c and d). Y axes have been truncated but range from -100 to +100.

Table S4: Summary of the relationship between recent stress (Perceived stress Scale: PSS) and temporal summation at the lumbar test site. * Temporal summation data at the lumbar site were missing for one participant due to a technical error with saving the data.

| Predictors | **Unadjusted model of main effect** | | | **Unadjusted model with sex interaction term** | | | **Adjusted model with sex interaction term** | | |
| --- | --- | --- | --- | --- | --- | --- | --- | --- | --- |
|  | Estimates | 95% CI | *p* | Estimates | 95% CI | *p* | Estimates | 95% CI | *p* |
| Intercept | 6.65 | -2.12; 15.41 | 0.135 | 7.93 | -6.39; 22.26 | 0.27 | 21.94 | 3.15; 40.73 | **0.02** |
| PSS score | 0.27 | -0.20; 0.74 | 0.257 | 0.03 | -0.81; 0.88 | 0.94 | 0.23 | -0.65; 1.11 | 0.60 |
| Sex (female) |  |  |  | -0.75 | -19.01; 17.52 | 0.94 | -1.12 | -19.72; 17.47 | 0.91 |
| PSS score x sex (female) |  |  |  | 0.28 | -0.74; 1.31 | 0.59 | 0.32 | -0.71; 1.36 | 0.54 |
| Age |  |  |  |  |  |  | -0.32 | -0.71; 0.08 | 0.11 |
| CTQ-SF total score |  |  |  |  |  |  | 0.02 | -0.13; 0.18 | 0.77 |
| PSQI score |  |  |  |  |  |  | -0.83 | -1.87; 0.21 | 0.12 |
| Diagnosis of major depressive disorder |  |  |  |  |  |  | 2.38 | -9.67; 16.24 | 0.62 |
| Recent COVID-19 infection |  |  |  |  |  |  | -0.43 | -13.39; 12.53 | 0.95 |
| Recent acute illness |  |  |  |  |  |  | -7.91 | -14.92; -0.90 | **0.03** |
| Diagnosis of chronic illness |  |  |  |  |  |  | 4.75 | -6.92; 16.43 | 0.42 |
| Observations* | 94 | | | 94 | | | 94 | | |
| R^2^ / R^2^ adjusted | 0.014/0.003 | | | 0.031/-0.002 | | | 0.125/0.020 | | |
| PSS = Perceived Stress Scale; CTQ-SF = Childhood Trauma Questionnaire-short form; PSQI = Pittsburgh Sleep Quality Index. | | | | | | | | | |

Table S5: Summary of the relationship between recent stress (Perceived stress Scale: PSS) and temporal summation at the deltoid test site.

| Predictors | **Unadjusted model of main effect** | | | **Unadjusted model with sex interaction term** | | | **Adjusted model with sex interaction term** | | |
| --- | --- | --- | --- | --- | --- | --- | --- | --- | --- |
|  | Estimates | 95% CI | *p* | Estimates | 95% CI | *p* | Estimates | 95% CI | *p* |
| Intercept | 6.81 | 0.31; 13.32 | **0.04** | 8.76 | -2.15; 19.67 | 0.11 | 16.61 | 1.96; 31.26 | **0.03** |
| PSS score | 0.19 | -0.16; 0.54 | 0.28 | 0.08 | -0.56; 0.72 | 0.81 | 0.19 | -0.49; 0.88 | 0.56 |
| Sex (female) |  |  |  | -3.05 | -16.79; 10.70 | 0.66 | -3.97 | -18.40; 10.45 | 0.59 |
| PSS score x sex (female) |  |  |  | 0.17 | -0.61; 0.94 | 0.67 | 0.21 | -0.60; 1.01 | 0.61 |
| Age |  |  |  |  |  |  | -0.15 | -0.45; 0.15 | 0.33 |
| CTQ-SF total score |  |  |  |  |  |  | -0.00 | -0.12; 0.12 | 0.97 |
| PSQI score |  |  |  |  |  |  | -0.52 | -1.33; 0.29 | 0.21 |
| Diagnosis of major depressive disorder |  |  |  |  |  |  | 5.16 | -4.93; 15.26 | 0.31 |
| Recent COVID-19 infection |  |  |  |  |  |  | -3.08 | -13.12; 6.97 | 0.54 |
| Recent acute illness |  |  |  |  |  |  | -3.35 | -8.81; 2.11 | 0.23 |
| Diagnosis of chronic illness |  |  |  |  |  |  | 2.42 | -6.33; 11.27 | 0.59 |
| Observations | 95 | | | 95 | | | 95 | | |
| R^2^ / R^2^ adjusted | 0.012/0.002 | | | 0.015/-0.018 | | | 0.068/-0.043 | | |
| PSS = Perceived Stress Scale; CTQ-SF = Childhood Trauma Questionnaire-short form; PSQI = Pittsburgh Sleep Quality Index. | | | | | | | | | |

###### d. Outcome: conditioned pain modulation

The unadjusted model satisfied the underlying assumptions of linear regression (Figs S14 and S15) and found no evidence of an association between PSS and CPM at either the lumbar (β=0.36 [95%CI: -0.41; 1.14], *p* = 0.53) or the deltoid site (β=-0.38 [95%CI: -0.99; 0.24], *p* = 0.32). When sex was included as an interaction term, both the unadjusted and the covariate-adjusted models found no evidence of an association between PSS and CPM at either the lumbar or the deltoid site, and there was no evidence of an effect of sex, or an effect of the interaction between PSS and sex (Fig S16 and Tables S6 and S7).

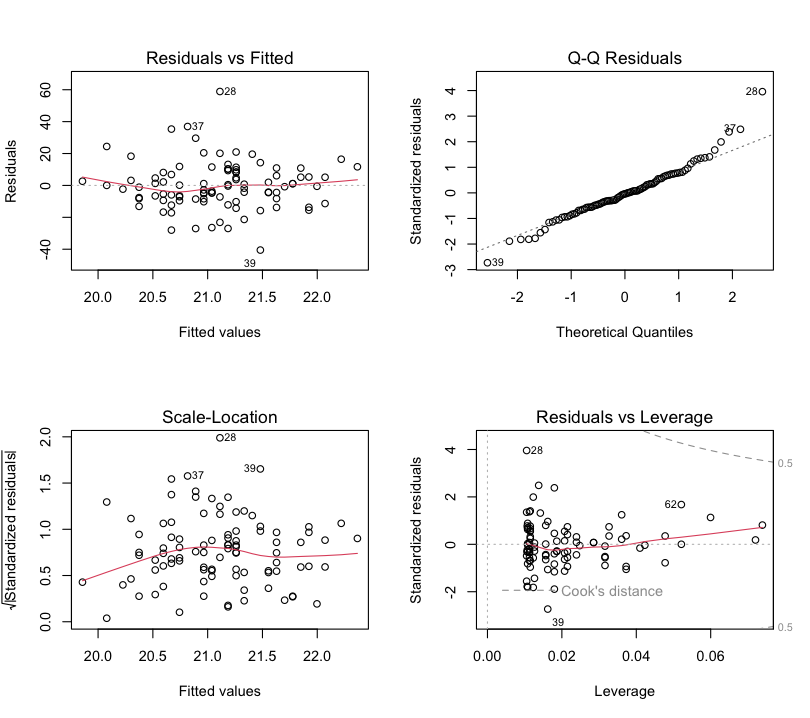

Figure S14: Plots of model assumptions for the conventional linear regression investigating the relationship between Perceived Stress Scale (PSS) and conditioned pain modulation at the lumbar site.

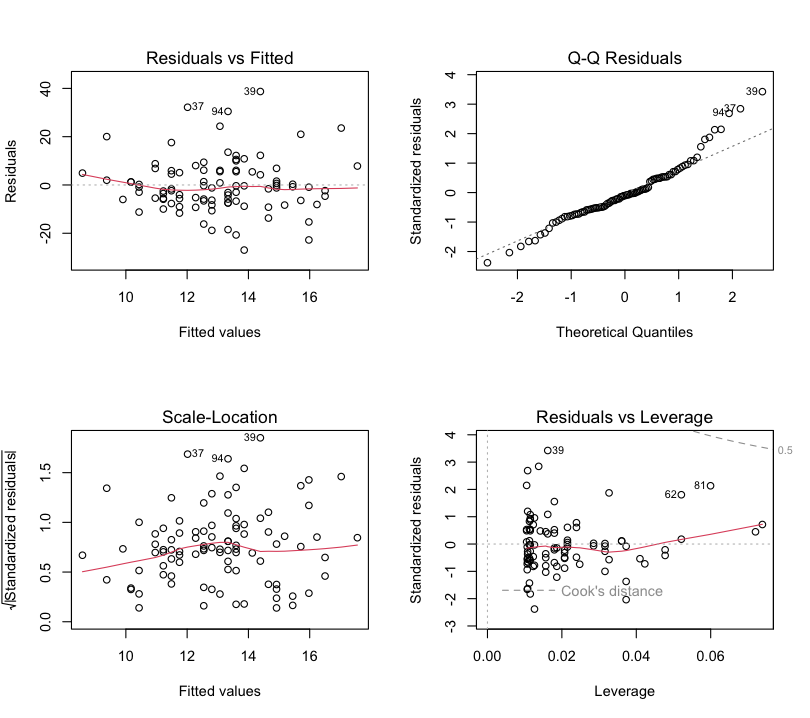

Figure S15: Plots of model assumptions for the conventional linear regression investigating the relationship between Perceived Stress Scale (PSS) and conditioned pain modulation at the deltoid site.

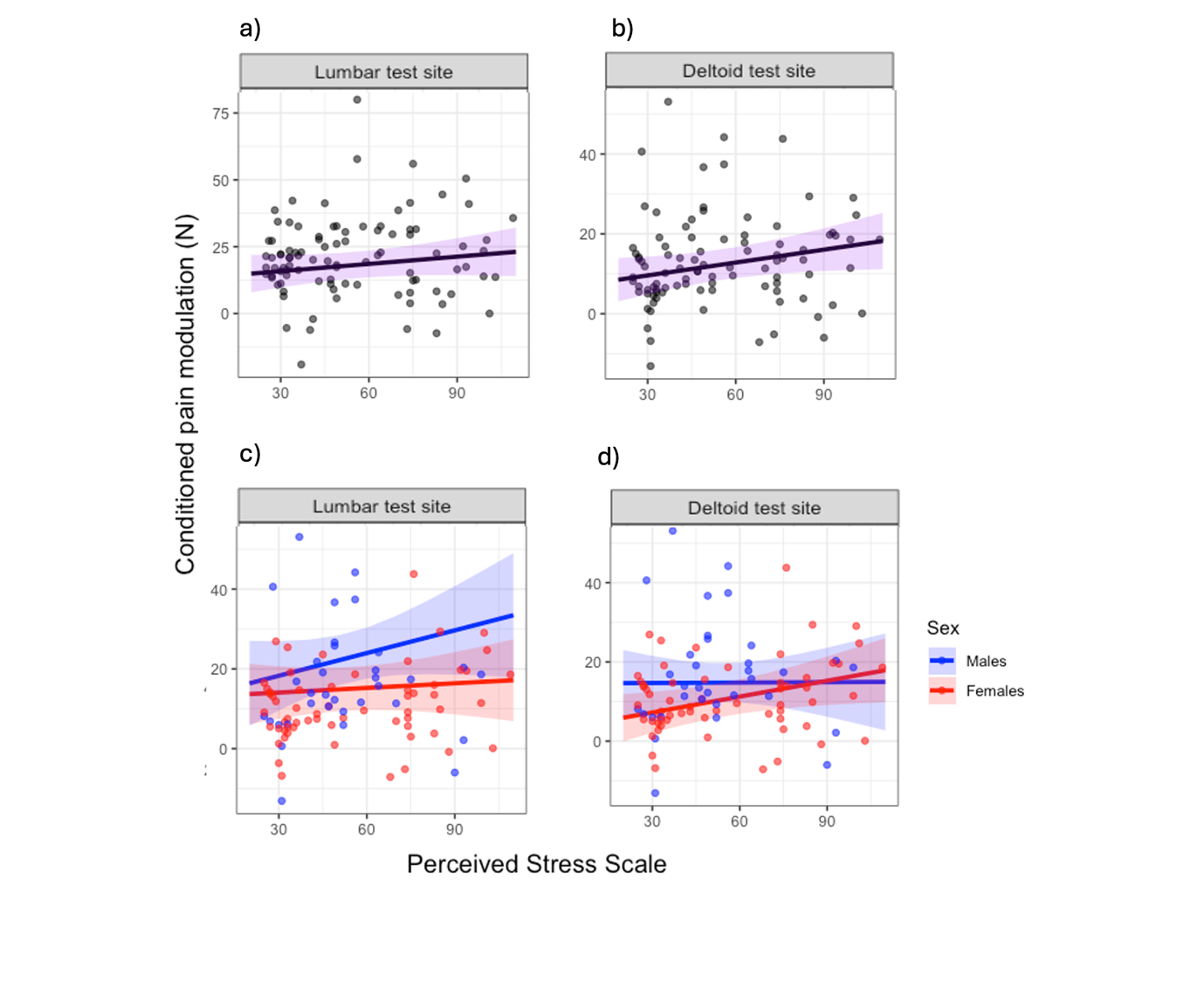

Figure S16: Plot of the association between the Perceived Stress Scale (PSS) and conditioned pain modulation. Plots show the observed values (dots) and the predicted regression line and 95% confidence interval (ribbon) for the covariate-adjusted model for the full cohort (plots a and b) and with sex as an interaction term (plots c and d).

Table S6: Summary of the relationship between recent stress (Perceived stress Scale: PSS) and conditioned pain modulation at the lumbar test site.

|  | **Unadjusted model of main effect** | | | **Unadjusted model with sex interaction term** | | | **Adjusted model with sex interaction term** | | |
| --- | --- | --- | --- | --- | --- | --- | --- | --- | --- |
| Predictors | Estimates | 95% CI | *p* | Estimates | 95% CI | *p* | Estimates | 95% CI | *p* |
| Intercept | 22.37 | 14.29; 30.44 | **<0.001** | 17.15 | 4.22; 30.08 | **0.01** | 0.74 | -15.77; 17.25 | 0.93 |
| PSS score | -0.07 | -0.51; 0.36 | 0.74 | 0.56 | -0.20; 1.33 | 0.15 | 0.36 | -0.41; 1.14 | 0.53 |
| Sex (female) |  |  |  | 5.66 | -10.63; 21.96 | 0.49 | 5.56 | 10.70; 21.82 | 0.50 |
| PSS score x sex (female) |  |  |  | -0.81 | -1.73; 0.11 | 0.08 | -0.80 | 1.70; 0.11 | 0.08 |
| Age |  |  |  |  |  |  | 0.29 | -0.06; 0.63 | 0.10 |
| CTQ-SF total score |  |  |  |  |  |  | 0.08 | -0.05; 0.22 | 0.24 |
| PSQI score |  |  |  |  |  |  | 0.55 | -0.36; 1.46 | 0.23 |
| Diagnosis of major depressive disorder |  |  |  |  |  |  | 5.62 | -5.77; 17.00 | 0.33 |
| Recent COVID-19 infection |  |  |  |  |  |  | 8.99 | -2.33; 20.31 | 0.12 |
| Recent acute illness |  |  |  |  |  |  | 5.78 | -0.37; 11.94 | 0.07 |
| Diagnosis of chronic illness |  |  |  |  |  |  | -8.73 | 18.71; 1.25 | 0.09 |
| Observations | 95 | | | 95 | | | 95 | | |
| R^2^ / R^2^ adjusted | 0.001/-0.010 | | | 0.092/0.062 | | | 0.223/0.130 | | |
| PSS = Perceived Stress Scale; CTQ-SF = Childhood Trauma Questionnaire-short form; PSQI = Pittsburgh Sleep Quality Index. | | | | | | | | | |

Table S7: Summary of the relationship between recent stress (Perceived stress Scale: PSS) and conditioned pain modulation at the deltoid test site.

|  | **Unadjusted model of main effect** | | | **Unadjusted model with sex interaction term** | | | **Adjusted model with sex interaction term** | | |
| --- | --- | --- | --- | --- | --- | --- | --- | --- | --- |
| Predictors | Estimates | 95% CI | *p* | Estimates | 95% CI | *p* | Estimates | 95% CI | *p* |
| Intercept | 17.56 | 11.40; 23.71 | **<0.001** | 21.42 | 11.35; 31.50 | **<0.001** | 16.07 | 2.85; 29.28 | **0.02** |
| PSS score | -0.26 | -0.60; 0.07 | 0.12 | -0.30 | -0.90; 0.29 | 0.31 | -0.38 | -0.99; 0.24 | 0.32 |
| Sex (female) |  |  |  | -7.41 | -20.10; 5.29 | 0.25 | -7.03 | -20.04; 5.99 | 0.29 |
| PSS score x sex (female) |  |  |  | 0.14 | -0.58; 0.85 | 0.70 | 0.15 | -0.57; 0.88 | 0.68 |
| Age |  |  |  |  |  |  | -0.01 | -0.28; 0.27 | 0.95 |
| CTQ-SF total score |  |  |  |  |  |  | 0.10 | -0.01; 0.21 | 0.07 |
| PSQI score |  |  |  |  |  |  | 0.00 | -0.73; 0.73 | 1.00 |
| Diagnosis of major depressive disorder |  |  |  |  |  |  | 4.54 | -4.58; 13.65 | 0.33 |
| Recent COVID-19 infection |  |  |  |  |  |  | 8.17 | -0.89; 17.23 | 0.08 |
| Recent acute illness |  |  |  |  |  |  | 0.80 | -4.13; 5.73 | 0.75 |
| Diagnosis of chronic illness |  |  |  |  |  |  | -3.94 | -11.93; 4.05 | 0.33 |
| Observations | 95 | | | 95 | | | 95 | | |
| R^2^ / R^2^ adjusted | 0.026/0.016 | | | 0.074/0.043 | | | 0.163/0.064 | | |
| PSS = Perceived Stress Scale; CTQ-SF = Childhood Trauma Questionnaire-short form; PSQI = Pittsburgh Sleep Quality Index. | | | | | | | | | |
